# Feasibility of a Cluster Randomised Trial on the Effect of Trauma Life Support Training: A Pilot Study

**DOI:** 10.1101/2024.03.13.24304236

**Authors:** Trauma life support training Effectiveness Research Network (TERN) collaborators, Martin Gerdin Wärnberg, Debojit Basak, Johanna Berg, Shamita Chatterjee, Li Felländer-Tsai, Geeta Ghag, Monty Khajanchi, Tamal Khan, Catherine Juillard, Vipul Nandu, Nobhojit Roy, Rajdeep Singh, Kapil Dev Soni, Lovisa Strömmer

**Affiliations:** Institute of Post Graduate Medical Education and Research, Kolkata, India; Department of Global Public Health, Karolinska Institutet, Stockholm, Sweden; Emergency Medicine, Department of Internal and Emergency Medicine, Skåne University Hospital, Malmö, Sweden; Department of Surgery, Institute of Post Graduate Medical Education and Research, Kolkata, India; Division of Orthopaedics and Biotechnology, Department of Clinical Science Intervention and Technology (CLINTEC), Karolinska Institutet, Stockholm, Sweden; Department of Reconstructive Orthopedics, Karolinska University Hospital, Stockholm, Sweden; Department of Surgery, HBT Medical College And Dr. R N Cooper Municipal General Hospital, Mumbai, India; WHO Collaboration Centre for Research in Surgical Care Delivery in LMIC, Mumbai, India; Seth G. S. Medical College and K.E.M. Hospital, Mumbai, India; Senior Research fellow, All India Institute of Medical Sciences, New Delhi, India; Division of General Surgery, Department of Surgery, David Geffen School of Medicine at UCLA, Los Angeles, California, United States of America; Program for Global Surgery and Trauma, The George Institute of Global Health, New Delhi, India; Department of Surgery, Maulana Azad Medical College, New Delhi, India; Critical and Intensive Care, JPN Apex Trauma Center, All India Institute of Medical Sciences, New Delhi, India; Division of Surgery and Oncology, Department of Clinical Science, Intervention and Technology (CLINTEC), and Department of Global Public Health, Karolinska Institutet, Stockholm, Sweden

**Keywords:** Trauma management, Accident and emergency medicine, Education and training

## Abstract

**Importance:** There is no high-quality evidence to show that trauma life support training programmes improve patient outcomes.

**Objective:** To assess the feasibility of conducting a cluster randomised controlled trial comparing the effect of Advanced Trauma Life Support^®^ (ATLS^®^) and Primary Trauma Care (PTC) with standard care on patient outcomes, and to estimate probable effect sizes and other measures needed for the sample size calculations of a full-scale trial.

**Design:** A pilot pragmatic three-armed parallel, cluster randomised, controlled trial between April 2022 and February 2023. Patient follow up was 30 days.

**Setting:** Tertiary care hospitals across metropolitan areas in India.

**Participants:** Adult trauma patients and residents managing these patients.

**Interventions:** ATLS^®^ or PTC training for residents in the intervention arms.

**Main Outcomes and Measures:** The outcomes were consent rate, lost to follow up rate, pass rate, missing data rates, differences in distribution between observed and data extracted from medical records as well as all cause and in-hospital mortality at 30 days from the time of arrival to the emergency department.

**Results:** Two hospitals were randomised to ATLS^®^, two to PTC, and three to standard care. We included 376 patients and 22 residents. The percentage of patients who consented to follow up was 77% and the percentage of residents who consented to training was 100%. The lost to follow up rate was 14%. The pass rate was 100%. The missing data was overall low for key variables. Data collected through observations were similar to data extracted from medical records, but there was more missing data in the extracted data. Twenty-two (16%) patients died within 30 days in the standard care arm, one (4%) patient in the ATLS^®^ arm, and three (5%) patients in the PTC arm.

**Conclusions and Relevance:** Conducting a full-scale cluster randomised controlled trial comparing the effects of ATLS^®^, PTC, and standard care on patient outcomes will be feasible, especially if such a trial would use data and outcomes available in medical records.

**Key Points:** 

**Question:** Is it feasible to conduct a cluster randomised trial comparing trauma life support training with standard care?

**Findings:** In this pilot cluster randomized trial that included 376 patients and 22 residents from seven hospitals, we found high consent rates, low lost to follow up rates, and low missing data for key variables.

**Meaning:** Conducting a full-scale cluster cluster trial comparing the effects of trauma life support training with standard care on patient outcomes will be feasible, especially if such a trial would use data and outcomes available in medical records.

## Introduction

Trauma, defined as the clinical entity composed of physical injury and the body’s associated response, causes 4.3 millions deaths every year^1^. Several trauma life support training programs have been developed to improve the early management of patients in hospital by providing a structured framework to assessment and treatment^2–4^.

The proprietary Advanced Trauma Life Support^®^ (ATLS^®^) and the low-cost alternative Primary Trauma Care (PTC) are two widely established trauma life support training programmes with over a million physicians trained in over 80 countries^5,6^. Observational studies indicate that these programmes may improve patient outcomes^7–19^, but there is no high quality evidence from controlled trials to support this^2–4,20–22^.

Several studies, including at least two randomised studies^23,24^, show that ATLS^®^ is associated with improved knowledge and skills among providers^2^. Observational evidence suggests that PTC also leads to improved provider skills^4^. The missing link is then whether these improved knowledge and skills translate into measurably improved patient outcomes.

Systematic reviews call for controlled trials in settings where these programmes are not routinely implemented^3,4^, because conducting such effectiveness trials in settings where they are part of the standard of care is not possible. Many settings without routinely implemented trauma life support training are in low-and middle income countries, where trial logistics can be more challenging.

We therefore aimed to assess the feasibility of conducting a cluster randomised controlled trial comparing ATLS^®^ and PTC with standard care, and to estimate probable effect sizes and other measures needed for the sample size calculations of a full-scale trial.

## Methods

### Trial Design

We piloted a three-armed cluster randomised controlled trial^25^. There were a standard care arm and two intervention arms, ATLS^®^ and PTC training. We collected data for four months in all three arms, first during a one month observation phase and then during a three month intervention phase (or continued observation in the standard care arm). This design allowed us to assess outcomes both as final values and as change from baseline.

### Study Setting

We conducted this pilot study in seven tertiary hospitals across metropolitan areas in India, where neither ATLS^®^, PTC, nor any other established trauma life support training program is routinely taught.

### Standard Care

Standard care varies across hospitals in India, but most surgical and emergency medicine departments in India organise their physicians in units. These units include both faculty members and residents, who are assigned a specific day of the week when they are posted in the emergency department. In the emergency department, trauma patients are initially assessed by residents who also resuscitate patients, perform interventions and refer patients for imaging or other investigations. Compared with other settings where a trauma team approach is adopted, nurses and other healthcare professionals are only involved to a limited extent during the initial management.

### Intervention

In each intervention arm the residents in one or two units were trained in either ATLS^®^ or PTC. For the purpose of this pilot study, our target was to train a minimum of 75% of residents in each unit. We did not train the units’ faculty, because they are typically not directly involved in the initial management of trauma patients. The ATLS^®^ training was conducted in an ATLS^®^ certified training centre in Mumbai, according to the standard ATLS^®^ curriculum^5^. The PTC training was conducted in New Delhi, according to the standard PTC curriculum^6^. We did not modify or adapt the delivery or content of these programmes during this pilot study.

### Eligibility Criteria for Cluster and Participants

#### Hospitals

We included tertiary care hospitals in metropolitan areas in India that admitted more than 400 adult patients with trauma each year, and that had operation theatres, X-ray, CT, and ultrasound facilities, and blood bank available around the clock.

#### Clusters

We defined a cluster as one or more units of physicians providing trauma care in the emergency department of Indian tertiary care hospitals. To be eligible, units could have no more than 25% of their physicians trained in either ATLS^®^, PTC, or similar training programs before the start of the pilot study. Those residents who had received training in the last five years were considered as trained. The figure of 25% was decided through consensus in the research team, to balance feasibility and contamination of results. The principal investigator at each hospital selected the units for training. We randomised on the hospital level to avoid contamination between intervention arms and the standard care arms.

#### Residents

We trained resident doctors doing their speciality training in surgery or emergency medicine managing trauma patients in the emergency department and who were expected to remain in the participating hospitals for at least one year from the time of the training.

Consent was sought from the residents in each of the intervention groups before they underwent the ATLS^®^ or PTC training.

#### Patients

We included persons who were 15 years or older and presented to the emergency department at participating hospitals with a history of trauma when a designated unit was on duty. History of trauma was defined as having any of the external causes of morbidity and mortality listed in block V01-Y36, chapter 20 of the International Classification of Disease version 10 (ICD-10) codebook as reason for presenting.

### Outcomes

We measured a large number of outcomes to help plan and assess the feasibility of a full scale trial. A list of outcomes is available in Supplementary Table S1. Our main outcomes were:

- Consent rate of patients and residents. This was equal to the percentage of patients or residents who consented to be included, out of the total number of eligible patients or residents.
- Lost to follow up rate. This applied only to patients and was equal to the percentage of patients who did not complete 30 day follow up, out of all enrolled patients.
- Pass rate. This applied only to residents in the intervention arms and was equal the percentage of residents who passed the training programme, out of the total number of trained residents.
- Missing data rate. This applied to each outcome and variable and was equal to the percentage of missing values.
- Differences in distributions of observed and extracted data. This applied to each outcome and variable and compared the distributions of data collected by observations versus extracted from hospital records.
- All cause and in-hospital mortality within 30 days from the time of arrival to the emergency department among patients.

### Participant Timeline and Inclusion

#### Patients

Arriving patients were screened for eligibility and consented, if conscious. Unconscious patients were consented by the patient’s representative. This proxy consent was reaffirmed by the patient, on regaining consciousness. We followed up patients at 24 hours after arrival at the emergency department, and up to 30 days after arrival at the emergency department.

#### Residents

Participating units were screened for eligibility once hospitals confirmed their participation. All residents in these units were approached to consent to training if their hospital was randomised to either of the intervention arms. The training was conducted approximately one month after the study started in that hospital.

### Sample size

We did not conduct a formal power calculation for this pilot study, as the purpose was to assess the feasibility of the trial logistics and research methods.

### Allocation and blinding

We used simple randomisation implemented using sealed envelopes to allocate sites to trial arms. We did not blind investigators, residents or patients to the intervention.

### Data Collection

Data was collected over a four-month period. A research officer collected data on all patients who presented on the days and shifts when participating residents were assigned to trauma care. The research officers observed care and interviewed residents and patients, and also extracted data from the hospital records. We followed up admitted patients for their complications and other in-hospital outcome measures. Patients who were not admitted or who were discharged before the end of the study were followed up telephonically for mortality outcomes and quality of life outcomes.

### Variables

The research officers collected data on demographics, vital signs, management details including imaging and surgery, and details of any injury sustained. All injuries were coded according to the International Classification of Diseases version 10 (ICD-10). Based on these ICD-10 codes, we calculated the Injury Severity Score using the R package `icdpicr`^26^. For a convenience sample of patients we also extracted data from medical records, to be able to compare the distribution of this data with the distribution of data collected through direct observations.

### Patient and public involvement

We conducted community consultations to collect inputs from patients, their caregivers, patient groups, and resident doctors to be used in the selection of outcome measures and implementation of the full-scale trial. The results of these consultations are published separately^27^.

### Data monitoring

We conducted weekly online meetings to monitor the study and data collection. We conducted one interim analysis approximately halfway through the study, and decided to complete the study as residents and patients were consenting to be included in the study and key variables including mortality outcomes could be collected. We did not use a data monitoring committee.

### Statistical Methods

We used the R version 4.4.1 (2024-06-14) Statistical Software for all analyses^28^. We analysed all data using descriptive statistics and did not perform any formal hypothesis tests^29^. Quantitative variables are summarised as median and interquartile range. Qualitative variables are presented as absolute numbers and percentages. We used an empty generalised linear mixed model to estimate the intracluster correlation coefficient.

We compared patients outcomes in all possible combinations of trial arms. In each combination we compared both differences in final values and differences in change from baseline. For the intervention arms the change from baseline was calculated as the difference between the one month period of data collection before the training was undertaken and the three month period after the training. For the control arm the data collection period was four months and the difference from baseline was calculated as the difference between the first one month and the following three months.

Within each combination of trial arms we had planned to conduct subgroup analyses of men, women, blunt multisystem trauma, penetrating trauma, shock (systolic blood pressure ≤ 90 mmHg), severe traumatic brain injury, and elderly (≥65 years)^30^. These subgroups were however too small to allow for meaningful analyses, and are therefore reported descriptively. We calculated both absolute and relative differences for each comparison, along with 75, 85, and 95% confidence intervals. We used an empirical bootstrap procedure with 1000 draws to estimate these confidence intervals.

In the interest of space, only the 95% confidence intervals are presented for all comparisons in Supplementary Tables. The remaining results are available from the corresponding author on request.

### Ethics and Dissemination

We were granted research ethics approval from the institutional ethics committees at each participating hospital. For each participating hospital, the approvals were HBTMC/266/SURGERY for Dr R N Cooper Municipal General Hospital in Mumbai, IEC(II)/OUT/134/2022 for Seth GS Medical College and KEM Hospital in Mumbai, ICC/214/22/20/05/2022 for Lokmanya Tilak Municipal Medical College and General Hospital, CREC/2022/FEB/1(ii) for MEDICA Superspeciality Hospital in Kolkata, MC/KOL/IEC/NON-SPON/1217/11/21 for Medical College, Kolkata, NRSMC/IEC/93/2021 for Nilratan Sircar Medical College & Hospital in Kolkata, and finally IEC-03/2022-2332 for the Postgraduate Institute of Medical Education and Research, Chandigarh.

## Results

We enrolled 376 trauma patients from seven participating centres between April 2022 and February 2023. The standard care arm enrolled 202 patients, the ATLS^®^ arm enrolled 44 patients, and the PTC arm enrolled 130 patients. We trained a total of 22 residents, seven in ATLS^®^, and 15 in PTC.

The study flow diagram is shown in Figure 1 and patient sample characteristics across trial arms are shown in Table 1. Overall, the number of females were 86 (23%), the median (IQR) age was 33 (24, 46) years, and the median ISS (IQR) was 4 (1, 8). A total of 32 (10%) patients died within 30 days after arrival to the emergency department, and 29 (8%) patients died in hospital. The intracluster correlation coefficients was 0.022 for 30-day mortality and 0.017 for in-hospital mortality.

**Table 1.**
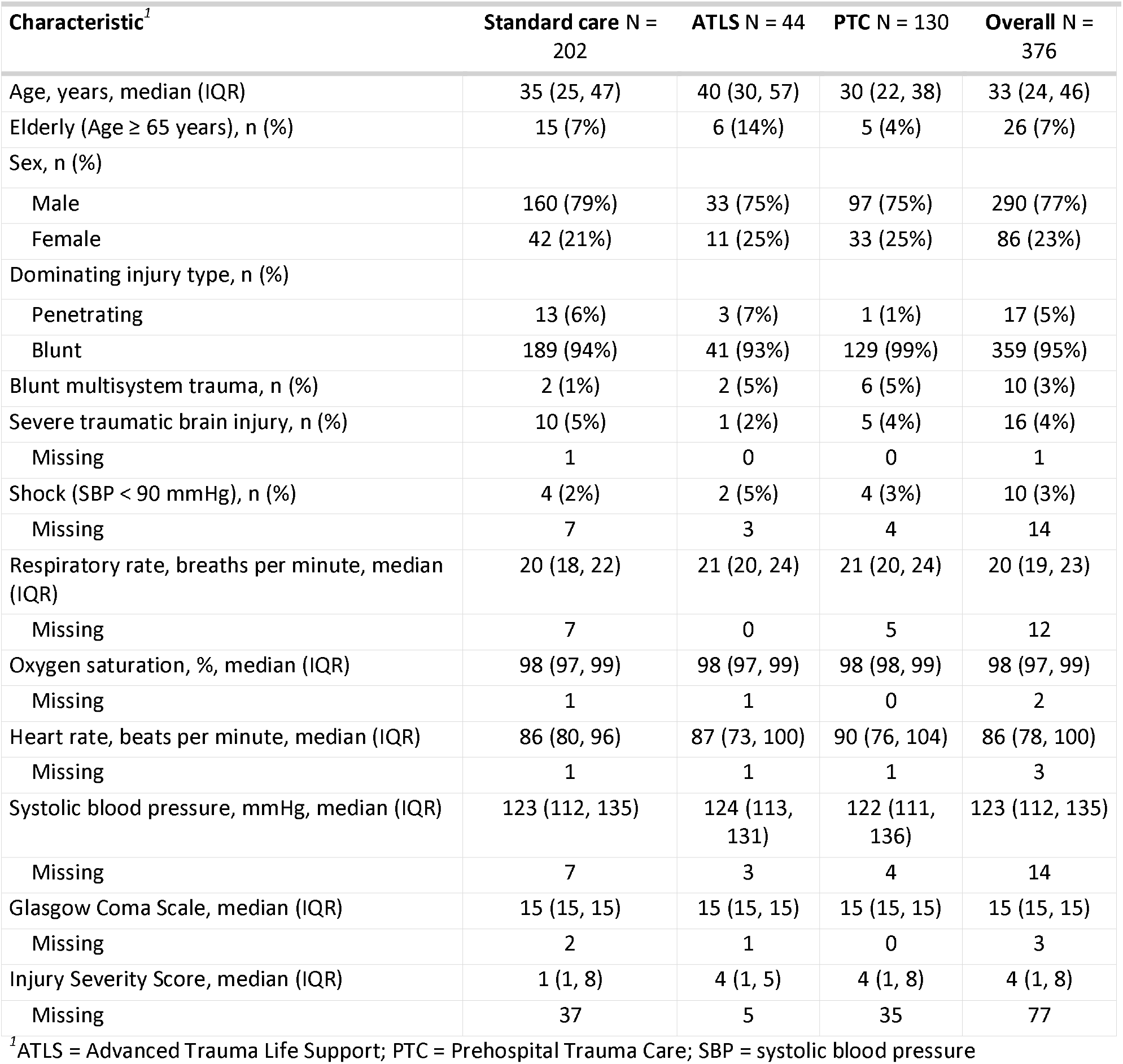
Patient sample characteristics.

| Characteristic<sup>1</sup> | Standard care N = 202 | ATLS N = 44 | PTC N = 130 | Overall N = 376 |
| --- | --- | --- | --- | --- |
| Age, years, median (IQR) | 35 (25, 47) | 40 (30, 57) | 30 (22, 38) | 33 (24, 46) |
| Elderly (Age ≥ 65 years), n (%) | 15 (7%) | 6 (14%) | 5 (4%) | 26 (7%) |
| Sex, n (%) |  |  |  |  |
| Male | 160 (79%) | 33 (75%) | 97 (75%) | 290 (77%) |
| Female | 42 (21%) | 11 (25%) | 33 (25%) | 86 (23%) |
| Dominating injury type, n (%) |  |  |  |  |
| Penetrating | 13 (6%) | 3 (7%) | 1 (1%) | 17 (5%) |
| Blunt | 189 (94%) | 41 (93%) | 129 (99%) | 359 (95%) |
| Blunt multisystem trauma, n (%) | 2 (1%) | 2 (5%) | 6 (5%) | 10 (3%) |
| Severe traumatic brain injury, n (%) | 10 (5%) | 1 (2%) | 5 (4%) | 16 (4%) |
| Missing | 1 | 0 | 0 | 1 |
| Shock (SBP < 90 mmHg), n (%) | 4 (2%) | 2 (5%) | 4 (3%) | 10 (3%) |
| Missing | 7 | 3 | 4 | 14 |
| Respiratory rate, breaths per minute, median (IQR) | 20 (18, 22) | 21 (20, 24) | 21 (20, 24) | 20 (19, 23) |
| Missing | 7 | 0 | 5 | 12 |
| Oxygen saturation, %, median (IQR) | 98 (97, 99) | 98 (97, 99) | 98 (98, 99) | 98 (97, 99) |
| Missing | 1 | 1 | 0 | 2 |
| Heart rate, beats per minute, median (IQR) | 86 (80, 96) | 87 (73, 100) | 90 (76, 104) | 86 (78, 100) |
| Missing | 1 | 1 | 1 | 3 |
| Systolic blood pressure, mmHg, median (IQR) | 123 (112, 135) | 124 (113, 131) | 122 (111, 136) | 123 (112, 135) |
| Missing | 7 | 3 | 4 | 14 |
| Glasgow Coma Scale, median (IQR) | 15 (15, 15) | 15 (15, 15) | 15 (15, 15) | 15 (15, 15) |
| Missing | 2 | 1 | 0 | 3 |
| Injury Severity Score, median (IQR) | 1 (1, 8) | 4 (1, 5) | 4 (1, 8) | 4 (1, 8) |
| Missing | 37 | 5 | 35 | 77 |

<sup>1</sup>ATLS = Advanced Trauma Life Support; PTC = Prehospital Trauma Care; SBP = systolic blood pressure

**Figure 1.**
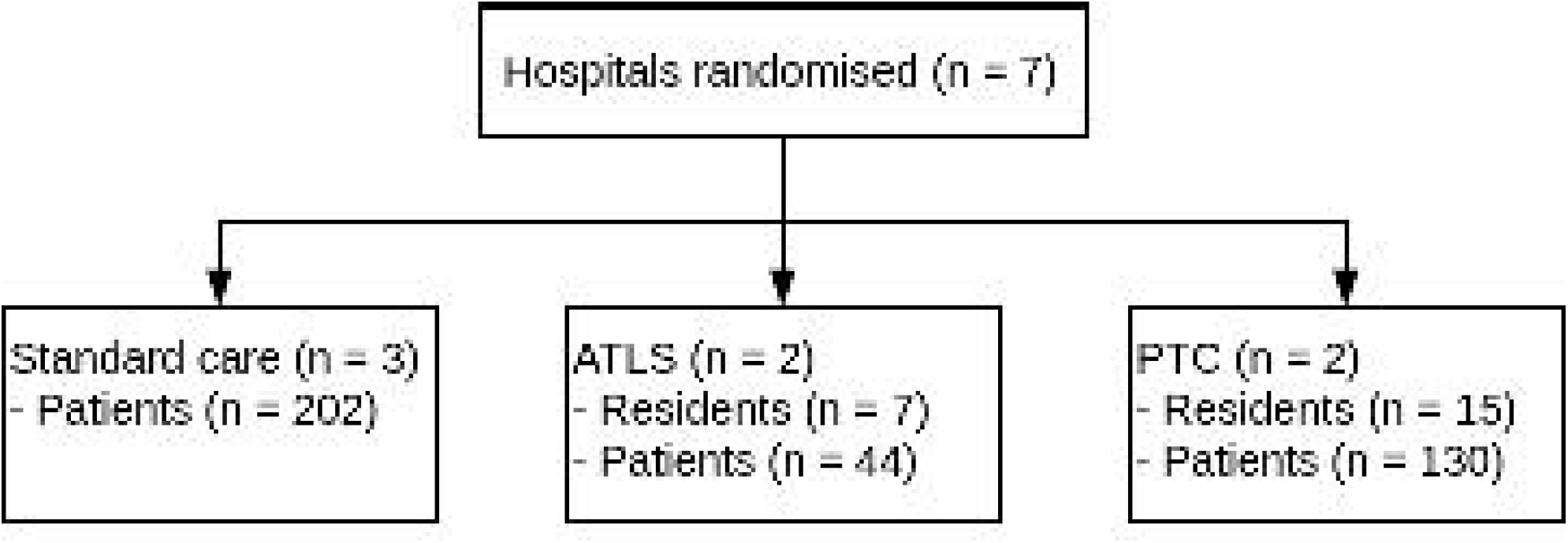
Study flow diagram. Abbreviations: ATLS, Advanced Trauma Life Support; PTC, Primary Trauma Care.

### Outcomes

The percentage of patients who consented to follow up was 77% and the percentage of residents who consented to training was 100%. Among patients, the lost to follow up rate was 14%. Among residents, the pass rate was 100%. The missing data rate ranged from 0 to 50%, with details for selected variables shown in Table 1 and in Supplementary Table S1. The variables with the maximum amount of missing data were in the cost of treatment, reported in Supplementary Tables S1-12. The differences in distributions between observed data and data extracted from medical records, for selected variables that were collected through observation or interview, are shown in Table 2. Overall, the data were similarly distributed, but there were considerably more missing values in the extracted data compared to the observed data.

**Table 2.** Differences in distributions between directly observed data and data extracted from medical records, for selected variables that were collected through observation or interview.

| Characteristic | Directly observed N = 55 | Medical records N = 55 |
| --- | --- | --- |
| Age, years, median (IQR) | 34 (27, 48) | 34 (25, 50) |
| Missing | 0 | 22 |
| Sex, n (%) |  |  |
| Female | 10 (18%) | 6 (18%) |
| Male | 45 (82%) | 27 (82%) |
| Missing | 0 | 22 |
| Dominating injury type, n (%) |  |  |
| Blunt | 52 (95%) | 29 (91%) |
| Penetrating | 3 (5%) | 3 (9%) |
| Missing | 0 | 23 |
| Respiratory rate, breaths per minute, median (IQR) | 21 (18, 24) | 18 (16, 20) |
| Missing | 0 | 37 |
| Oxygen saturation, %, median (IQR) | 98 (98, 99) | 98 (97, 100) |
| Missing | 0 | 29 |
| Heart rate, beats per minute, median (IQR) | 85 (80, 98) | 87 (84, 93) |
| Missing | 0 | 19 |
| Systolic blood pressure, mmHg, median (IQR) | 123 (112, 136) | 118 (110, 128) |
| Missing | 1 | 18 |

After training, a total of 22 (16%) patients in the standard care arm died within 30 days, compared to 1 (4%) patients in the ATLS^®^ arm and 3 (5%) patients in the PTC arm. The corresponding figures for in-hospital mortality were 19 (12%)%, 1 (4%)%, and 3 (4%)% for the standard care, ATLS^®^ and PTC arms respectively, as shown in Table 3. Overall, both in-hospital and 30-day mortality were substantially lower in the ATLS^®^ and PTC arms compared to the standard care arm, but the absolute numbers of deaths in the ATLS^®^ and PTC arms were very small. The results for all other outcomes are shown in Supplementary Tables S1-12.

**Table 3.** Mortality after training by the trial arms standard care, Advanced Trauma Life Support (ATLS) and Primary Trauma Care (PTC)

| Outcome | Arms |  |  | Differences |  |  |
| --- | --- | --- | --- | --- | --- | --- |
|  | Standard care N = 161<sup>1</sup> | ATLS N = 28<sup>1</sup> | PTC N = 73<sup>1</sup> | Standard care vs. ATLS | Standard care vs. PTC | ATLS vs. PTC |
| 30 day mortality | 22 (16%) | 1 (3.8%) | 3 (4.9%) | 12% | 11% | -1.1% |
| Unknown | 26 | 2 | 12 |  |  |  |
| In-hospital mortality | 19 (12%) | 1 (3.7%) | 3 (4.1%) | 8.2% | 7.8% | -0.41% |
| Unknown | 2 | 1 | 0 |  |  |  |

<sup>1</sup>n (%)

## Discussion

We show that it is feasible to conduct and collect data for a cluster randomized controlled trial comparing ATLS^®^ with PTC and standard care. Missing data were low for key variables, including the primary outcome and many secondary outcomes. Some variables, especially the cost of treatment (reported in Supplementary Table S1-12) had very high missing data rates and may not be feasible to include in a full-scale trial, or require different data collection methods. The missing data was substantially higher when data was extracted from medical records instead of being directly observed, but the data were similarly distributed, indicating that data collected from medical records is reliable even if it is less complete.

We found that the ATLS^®^ and PTC arms had lower 30-day mortality compared to the PTC and standard care arms. This finding could hint towards a potential effect of training physicians in trauma life support, but it is important to note that this pilot study was not powered to detect any differences in outcomes. The arms differed considerably in sample size, with the ATLS^®^ arm having the smallest sample size. This difference most likely 10 resulted from the randomisation process with a small number of heterogeneous clusters, and this heterogeneity highlights the importance of taking varying cluster sizes into account in the design of the full scale trial.

All-cause 30-day mortality was missing in 14% of patients. This may appear high, especially compared to for example the CRASH-2 and REACT-2 trials, which report missing primary outcome in less than 0.01% of patients^31,32^. Like many other trauma trials, both CRASH-2 and REACT-2 used in-hospital mortality as their primary outcome measure, whereas we attempted to follow up patients after discharge. Our missing data rate for in-hospital mortality was only 1%, which is comparable to previous trials.

During the course of this pilot we deviated from the protocol in several ways, and provide a detailed list as Supplementary material S13. Some key limitations of this pilot and therefore lessons to be learned and factored into the design of the full-scale trial include the lower than expected enrolment rates of some centres, centre specific management routines, and difficulties in collecting data on complications and cause of death.

We attempted to minimse the impact of the lower than expected enrolment rates by including a seventh centre, but careful assessments of patient volumes as part of the screening process will be needed for the full-scale trial. We decided to be pragmatic in selecting which residents to train and how to structure the data collection depending on how and by whom patients were initially managed, but this flexibility will need to be built into the full-scale trial protocol. Finally, we found that data on complications and cause of death were hard to identify and therefore the full-scale trial will need to include longer training of research officers if this data is to be collected.

Previous studies on the effect of ATLS^®^ or PTC training on patient outcomes are observational or quasi-experimental without a control group, with heterogeneous results^8^. Most studies have found that these programmes are associated with improved outcomes, although not all studies have found significant effects^7,9,10,12,14–18^. In contrast, some studies have found that these programmes may be associated with increased mortality^13,19^.

Considering the widespread use of trauma life support training, several systematic reviews call for trials in settings where these programmes are not routinely implemented^2–4^. Our study represents the first published attempt at a controlled trial of the effect of trauma life support training och patient outcomes, and we conclude that conducting a full-scale cluster randomised trial should be feasible after incorporating the lessons of this pilot.

## Data Availability

All data produced in the present study are available upon reasonable request to the authors.

## Contributorship statement

MGW conceived of the study, performed the analysis and drafted and revised the manuscript. AG, AM, CJ, DKV, HS, JB, KDS, LFT, LS, MH, MK, NR, PB, PP, RS, SD, and VK contributed to the design of the study. MGV, DKV, KDS, and MK drafted the first version of the protocol. AG, HS, and SD drafted the first version of the patient and public involvement activities. JB and PP drafted the first versions of the data management sections and wrote the data management plan. PB and PP drafted the first versions of the statistical analysis section. AG, AM, CJ, DKV, HS, JB, KDS, LFT, LS, MH, MGW, MK, NR, PB, PP, RS, SC, SD, and VK contributed to the refinement of the protocol. DB, JB, SC, LFT, GG, MK, TK, CJ, NR, RS, KDS, LS and VP interpreted the results and revised the manuscript. AR, AC, C, DK, GG, MK, MT, VK and VP are representatives of participating hospitals.

## Data Sharing Statement

The code for analysis is released publicly. The final anonymized dataset is available from the corresponding author on request.

