## Supplementary Tables for "Feasibility of a Cluster Randomised Trial on the Effect of Trauma Life Support Training: A Pilot Study"

Data collection from records ... 44
Selection of units for training ... 44
Timing of resident consent ... 44
Analysis level of feasibility outcomes ... 45

### Table S1. Extended sample characteristics

| Characteristic | Before training | | | | After training | | | | Overall |
|---|---|---|---|---|---|---|---|---|---|
| | **Standard care** N = 41[1] | **ATLS** N = 16[1] | **PTC** N = 57[1] | **Overall** N = 114[1] | **Standard care** N = 161[1] | **ATLS** N = 28[1] | **PTC** N = 73[1] | **Overall** N = 262[1] | **N = 376**[1] |
| **Age, years** | 32 (23, 46) | 46 (30, 61) | 30 (22, 38) | 33 (23, 46) | 35 (26, 47) | 37 (30, 55) | 30 (22, 38) | 34 (25, 45) | 33 (24, 46) |
| **Elderly (Age ≥ 65 years)** | 3 (7%) | 3 (19%) | 3 (5%) | 9 (8%) | 12 (7%) | 3 (11%) | 2 (3%) | 17 (6%) | 26 (7%) |
| **Sex** | | | | | | | | | |
| Male | 36 (88%) | 10 (63%) | 44 (77%) | 90 (79%) | 124 (77%) | 23 (82%) | 53 (73%) | 200 (76%) | 290 (77%) |
| Female | 5 (12%) | 6 (38%) | 13 (23%) | 24 (21%) | 37 (23%) | 5 (18%) | 20 (27%) | 62 (24%) | 86 (23%) |
| **Dominating injury type** | | | | | | | | | |
| Penetrating | 3 (7%) | 2 (13%) | 0 (0%) | 5 (4%) | 10 (6%) | 1 (4%) | 1 (1%) | 12 (5%) | 17 (5%) |
| Blunt | 38 (93%) | 14 (88%) | 57 (100%) | 109 (96%) | 151 (94%) | 27 (96%) | 72 (99%) | 250 (95%) | 359 (95%) |
| **Blunt multisystem trauma** | 0 (0%) | 1 (6%) | 3 (5%) | 4 (4%) | 2 (1%) | 1 (4%) | 3 (4%) | 6 (2%) | 10 (3%) |
| **Severe traumatic brain injury** | 3 (8%) | 0 (0%) | 3 (5%) | 6 (5%) | 7 (4%) | 1 (4%) | 2 (3%) | 10 (4%) | 16 (4%) |
| Missing | 1 | 0 | 0 | 1 | | | | | 1 |
| **Shock (SBP < 90 mmHg)** | 0 (0%) | 1 (7%) | 0 (0%) | 1 (1%) | 4 (3%) | 1 (4%) | 4 (6%) | 9 (4%) | 10 (3%) |
| Missing | 3 | 1 | 1 | 5 | 4 | 2 | 3 | 9 | 14 |
| **Respiratory rate, breaths per minute** | 20 (18, 21) | 22 (20, 24) | 21 (19, 23) | 20 (18, 22) | 20 (18, 22) | 21 (19, 24) | 22 (20, 25) | 21 (19, 23) | 20 (19, 23) |
| Missing | 4 | 0 | 3 | 7 | 3 | 0 | 2 | 5 | 12 |
| **Oxygen saturation, %** | 98 (98, 99) | 98 (96, 99) | 98 (97, 98) | 98 (97, 99) | 98 (97, 99) | 98 (97, 99) | 98 (98, 99) | 98 (98, 99) | 98 (97, 99) |
| Missing | 1 | 1 | 0 | 2 | | | | | 2 |
| **Heart rate, beats per minute** | 86 (80, 97) | 94 (74, 106) | 90 (79, 104) | 88 (80, 100) | 85 (80, 95) | 86 (73, 97) | 90 (74, 105) | 86 (78, 100) | 86 (78, 100) |
| Missing | 1 | 0 | 1 | 2 | 0 | 1 | 0 | 1 | 3 |
| **Systolic blood pressure, mmHg** | 126 (116, 130) | 128 (113, 150) | 123 (115, 138) | 124 (115, 133) | 123 (112, 136) | 124 (113, 130) | 120 (110, 136) | 123 (111, 136) | 123 (112, 135) |
| Missing | 3 | 1 | 1 | 5 | 4 | 2 | 3 | 9 | 14 |
| **Glasgow Coma Scale** | 15 (15, 15) | 15 (15, 15) | 15 (15, 15) | 15 (15, 15) | 15 (15, 15) | 15 (15, 15) | 15 (15, 15) | 15 (15, 15) | 15 (15, 15) |
| Missing | 1 | 1 | 0 | 2 | 1 | 0 | 0 | 1 | 3 |
| **Injury Severity Score** | 1 (1, 4) | 4 (1, 25) | 4 (2, 16) | 4 (1, 14) | 4 (1, 9) | 4 (1, 4) | 4 (1, 5) | 4 (1, 6) | 4 (1, 8) |

| Characteristic | Before training | | | | After training | | | | Overall |
|---|---|---|---|---|---|---|---|---|---|
| | **Standard care** N = 41[1] | **ATLS** N = 16[1] | **PTC** N = 57[1] | **Overall** N = 114[1] | **Standard care** N = 161[1] | **ATLS** N = 28[1] | **PTC** N = 73[1] | **Overall** N = 262[1] | **N = 376**[1] |
| Missing | 5 | 2 | 18 | 25 | 32 | 3 | 17 | 52 | 77 |
| **In-hospital mortality** | 2 (5%) | 0 (0%) | 4 (7%) | 6 (5%) | 19 (12%) | 1 (4%) | 3 (4%) | 23 (9%) | 29 (8%) |
| Missing | | | | | 2 | 1 | 0 | 3 | 3 |
| **30 day mortality** | 1 (3%) | 0 (0%) | 5 (10%) | 6 (6%) | 22 (16%) | 1 (4%) | 3 (5%) | 26 (12%) | 32 (10%) |
| Missing | 3 | 2 | 8 | 13 | 26 | 2 | 12 | 40 | 53 |
| **24 hour mortality** | 1 (2%) | 0 (0%) | 0 (0%) | 1 (1%) | 0 (0%) | 0 (0%) | 0 (0%) | 0 (0%) | 1 (0%) |
| Missing | 0 | 2 | 5 | 7 | 7 | 2 | 8 | 17 | 24 |
| **Self-ambulatory at discharge** | 37 (95%) | 11 (92%) | 47 (96%) | 95 (95%) | 135 (99%) | 24 (100%) | 68 (97%) | 227 (98%) | 322 (97%) |
| Missing | 2 | 4 | 8 | 14 | 24 | 4 | 3 | 31 | 45 |
| **Return to work** | 12 (41%) | 5 (83%) | 31 (74%) | 48 (62%) | 56 (53%) | 11 (58%) | 40 (70%) | 107 (59%) | 155 (60%) |
| Missing | 12 | 10 | 15 | 37 | 56 | 9 | 16 | 81 | 118 |
| **Pulmonary complication** | 0 (0%) | 0 (0%) | 1 (2%) | 1 (1%) | 0 (0%) | 0 (0%) | 0 (0%) | 0 (0%) | 1 (0%) |
| Missing | 2 | 6 | 11 | 19 | 33 | 5 | 10 | 48 | 67 |
| **Septic complication** | 1 (3%) | 0 (0%) | 2 (4%) | 3 (3%) | 1 (1%) | 0 (0%) | 2 (3%) | 3 (1%) | 6 (2%) |
| Missing | 1 | 6 | 12 | 19 | 33 | 5 | 10 | 48 | 67 |
| **Renal failure** | | | | | | | | | |
| No | 40 (100%) | 11 (100%) | 46 (100%) | 97 (100%) | 129 (100%) | 23 (100%) | 63 (100%) | 215 (100%) | 312 (100%) |
| Missing | 1 | 5 | 11 | 17 | 32 | 5 | 10 | 47 | 64 |
| **Coagulopathy** | 0 (0%) | 0 (0%) | 0 (0%) | 0 (0%) | 0 (0%) | 1 (4%) | 0 (0%) | 1 (0%) | 1 (0%) |
| Missing | 1 | 6 | 12 | 19 | 32 | 5 | 10 | 47 | 66 |
| **Need for reexploration or resurgery** | 0 (0%) | 0 (0%) | 1 (2%) | 1 (1%) | 3 (3%) | 0 (0%) | 1 (2%) | 4 (2%) | 5 (2%) |
| Missing | 1 | 4 | 10 | 15 | 42 | 6 | 10 | 58 | 73 |
| **Failure of conservative management** | 0 (0%) | 0 (0%) | 1 (2%) | 1 (1%) | 2 (2%) | 2 (8%) | 1 (2%) | 5 (2%) | 6 (2%) |
| Missing | 2 | 10 | 11 | 23 | 44 | 3 | 12 | 59 | 82 |
| **EQ-5D mobility at discharge** | | | | | | | | | |
| I have no problems in walking | 24 (62%) | 8 (67%) | 34 (83%) | 66 (72%) | 73 (62%) | 16 (89%) | 55 (92%) | 144 (73%) | 210 (73%) |
| I have some problems in walking | 7 (18%) | 1 (8%) | 5 (12%) | 13 (14%) | 22 (19%) | 2 (11%) | 3 (5%) | 27 (14%) | 40 (14%) |

| Characteristic | Before training | | | | After training | | | | Overall |
|---|---|---|---|---|---|---|---|---|---|
| | **Standard care** N = 41[1] | **ATLS** N = 16[1] | **PTC** N = 57[1] | **Overall** N = 114[1] | **Standard care** N = 161[1] | **ATLS** N = 28[1] | **PTC** N = 73[1] | **Overall** N = 262[1] | **N = 376**[1] |
| I am confined to bed | 8 (21%) | 3 (25%) | 2 (5%) | 13 (14%) | 23 (19%) | 0 (0%) | 2 (3%) | 25 (13%) | 38 (13%) |
| Missing | 2 | 4 | 16 | 22 | 43 | 10 | 13 | 66 | 88 |
| **EQ-5D self-care at discharge** | | | | | | | | | |
| I have no problems with self-care | 23 (59%) | 7 (58%) | 33 (80%) | 63 (68%) | 59 (50%) | 12 (67%) | 51 (85%) | 122 (62%) | 185 (64%) |
| I have some problems bathing or dressing myself | 6 (15%) | 2 (17%) | 5 (12%) | 13 (14%) | 36 (31%) | 2 (11%) | 5 (8%) | 43 (22%) | 56 (19%) |
| I am unable to bathe or dress myself | 10 (26%) | 3 (25%) | 3 (7%) | 16 (17%) | 23 (19%) | 4 (22%) | 4 (7%) | 31 (16%) | 47 (16%) |
| Missing | 2 | 4 | 16 | 22 | 43 | 10 | 13 | 66 | 88 |
| **EQ-5D usual activities at discharge** | | | | | | | | | |
| I have no problems in performing my usual activities | 21 (54%) | 6 (50%) | 31 (76%) | 58 (63%) | 55 (47%) | 11 (61%) | 51 (85%) | 117 (60%) | 175 (61%) |
| I have some problems in performing my usual activities | 11 (28%) | 3 (25%) | 6 (15%) | 20 (22%) | 45 (38%) | 4 (22%) | 5 (8%) | 54 (28%) | 74 (26%) |
| I am unable to perform my usual activities | 7 (18%) | 3 (25%) | 4 (10%) | 14 (15%) | 18 (15%) | 3 (17%) | 4 (7%) | 25 (13%) | 39 (14%) |
| Missing | 2 | 4 | 16 | 22 | 43 | 10 | 13 | 66 | 88 |
| **EQ-5D pain/discomfort at discharge** | | | | | | | | | |
| I have no pain or discomfort | 13 (33%) | 4 (33%) | 14 (34%) | 31 (34%) | 30 (25%) | 9 (50%) | 39 (65%) | 78 (40%) | 109 (38%) |
| I have moderate pain or discomfort | 16 (41%) | 8 (67%) | 26 (63%) | 50 (54%) | 41 (35%) | 9 (50%) | 18 (30%) | 68 (35%) | 118 (41%) |
| I have extreme pain or discomfort | 10 (26%) | 0 (0%) | 1 (2%) | 11 (12%) | 47 (40%) | 0 (0%) | 3 (5%) | 50 (26%) | 61 (21%) |
| Missing | 2 | 4 | 16 | 22 | 43 | 10 | 13 | 66 | 88 |
| **EQ-5D anxiety/depression at discharge** | | | | | | | | | |
| I am not anxious or depressed | 31 (79%) | 6 (50%) | 30 (73%) | 67 (73%) | 108 (92%) | 15 (83%) | 57 (95%) | 180 (92%) | 247 (86%) |
| I am moderately anxious or depressed | 7 (18%) | 6 (50%) | 10 (24%) | 23 (25%) | 8 (7%) | 3 (17%) | 3 (5%) | 14 (7%) | 37 (13%) |

| Characteristic | Before training | | | | After training | | | | Overall |
|---|---|---|---|---|---|---|---|---|---|
| | **Standard care** N = 41[1] | **ATLS** N = 16[1] | **PTC** N = 57[1] | **Overall** N = 114[1] | **Standard care** N = 161[1] | **ATLS** N = 28[1] | **PTC** N = 73[1] | **Overall** N = 262[1] | **N = 376**[1] |
| I am extremely anxious or depressed | 1 (3%) | 0 (0%) | 1 (2%) | 2 (2%) | 2 (2%) | 0 (0%) | 0 (0%) | 2 (1%) | 4 (1%) |
| Missing | 2 | 4 | 16 | 22 | 43 | 10 | 13 | 66 | 88 |
| **EQ-5D mobility at 30 day follow-up** | | | | | | | | | |
| I have no problems in walking | 20 (65%) | 4 (67%) | 39 (91%) | 63 (79%) | 79 (77%) | 16 (80%) | 57 (98%) | 152 (84%) | 215 (83%) |
| I have some problems in walking | 4 (13%) | 1 (17%) | 3 (7%) | 8 (10%) | 17 (17%) | 4 (20%) | 0 (0%) | 21 (12%) | 29 (11%) |
| I am confined to bed | 7 (23%) | 1 (17%) | 1 (2%) | 9 (11%) | 6 (6%) | 0 (0%) | 1 (2%) | 7 (4%) | 16 (6%) |
| Missing | 10 | 10 | 14 | 34 | 59 | 8 | 15 | 82 | 116 |
| **EQ-5D self-care at 30 day follow-up** | | | | | | | | | |
| I have no problems with self-care | 22 (71%) | 5 (83%) | 39 (91%) | 66 (83%) | 81 (79%) | 16 (80%) | 56 (97%) | 153 (85%) | 219 (84%) |
| I have some problems bathing or dressing myself | 2 (6%) | 1 (17%) | 2 (5%) | 5 (6%) | 15 (15%) | 2 (10%) | 0 (0%) | 17 (9%) | 22 (8%) |
| I am unable to bathe or dress myself | 7 (23%) | 0 (0%) | 2 (5%) | 9 (11%) | 6 (6%) | 2 (10%) | 2 (3%) | 10 (6%) | 19 (7%) |
| Missing | 10 | 10 | 14 | 34 | 59 | 8 | 15 | 82 | 116 |
| **EQ-5D usual activities at 30 day follow-up** | | | | | | | | | |
| I have no problems in performing my usual activities | 22 (71%) | 6 (100%) | 39 (91%) | 67 (84%) | 83 (81%) | 15 (75%) | 55 (95%) | 153 (85%) | 220 (85%) |
| I have some problems in performing my usual activities | 6 (19%) | 0 (0%) | 2 (5%) | 8 (10%) | 12 (12%) | 3 (15%) | 1 (2%) | 16 (9%) | 24 (9%) |
| I am unable to perform my usual activities | 3 (10%) | 0 (0%) | 2 (5%) | 5 (6%) | 7 (7%) | 2 (10%) | 2 (3%) | 11 (6%) | 16 (6%) |
| Missing | 10 | 10 | 14 | 34 | 59 | 8 | 15 | 82 | 116 |
| **EQ-5D pain/discomfort at 30 day follow-up** | | | | | | | | | |
| I have no pain or discomfort | 23 (74%) | 4 (67%) | 33 (77%) | 60 (75%) | 81 (79%) | 16 (80%) | 50 (86%) | 147 (82%) | 207 (80%) |
| I have moderate pain or discomfort | 8 (26%) | 2 (33%) | 9 (21%) | 19 (24%) | 19 (19%) | 4 (20%) | 8 (14%) | 31 (17%) | 50 (19%) |

| Characteristic | Before training | | | | After training | | | | Overall |
|---|---|---|---|---|---|---|---|---|---|
| | **Standard care** N = 41[1] | **ATLS** N = 16[1] | **PTC** N = 57[1] | **Overall** N = 114[1] | **Standard care** N = 161[1] | **ATLS** N = 28[1] | **PTC** N = 73[1] | **Overall** N = 262[1] | **N = 376**[1] |
| I have extreme pain or discomfort | 0 (0%) | 0 (0%) | 1 (2%) | 1 (1%) | 2 (2%) | 0 (0%) | 0 (0%) | 2 (1%) | 3 (1%) |
| Missing | 10 | 10 | 14 | 34 | 59 | 8 | 15 | 82 | 116 |
| **EQ-5D anxiety/depression at 30 day follow-up** | | | | | | | | | |
| I am not anxious or depressed | 30 (97%) | 6 (100%) | 37 (86%) | 73 (91%) | 97 (95%) | 17 (85%) | 56 (97%) | 170 (94%) | 243 (93%) |
| I am moderately anxious or depressed | 1 (3%) | 0 (0%) | 5 (12%) | 6 (8%) | 4 (4%) | 3 (15%) | 2 (3%) | 9 (5%) | 15 (6%) |
| I am extremely anxious or depressed | 0 (0%) | 0 (0%) | 1 (2%) | 1 (1%) | 1 (1%) | 0 (0%) | 0 (0%) | 1 (1%) | 2 (1%) |
| Missing | 10 | 10 | 14 | 34 | 59 | 8 | 15 | 82 | 116 |
| **Patient satisfaction** | | | | | | | | | |
| Very satisfied | 26 (67%) | 9 (64%) | 37 (73%) | 72 (69%) | 97 (70%) | 19 (79%) | 66 (94%) | 182 (78%) | 254 (76%) |
| Somewhat satisfied | 9 (23%) | 5 (36%) | 8 (16%) | 22 (21%) | 20 (14%) | 3 (13%) | 4 (6%) | 27 (12%) | 49 (15%) |
| Somewhat dissatisfied | 3 (8%) | 0 (0%) | 2 (4%) | 5 (5%) | 13 (9%) | 1 (4%) | 0 (0%) | 14 (6%) | 19 (6%) |
| Very dissatisfied | 1 (3%) | 0 (0%) | 4 (8%) | 5 (5%) | 8 (6%) | 1 (4%) | 0 (0%) | 9 (4%) | 14 (4%) |
| Missing | 2 | 2 | 6 | 10 | 23 | 4 | 3 | 30 | 40 |
| **Number of hospitalizations for this injury** | 0 (0%) | 1 (17%) | 3 (6%) | 4 (4%) | 6 (5%) | 0 (0%) | 0 (0%) | 6 (3%) | 10 (3%) |
| Missing | 2 | 10 | 8 | 20 | 34 | 3 | 11 | 48 | 68 |
| **EQ-5D health state at discharge** | 50 (3, 90) | 60 (40, 90) | 80 (60, 90) | 75 (40, 90) | 50 (4, 80) | 65 (50, 80) | 90 (79, 100) | 70 (5, 90) | 70 (10, 90) |
| Missing | 10 | 5 | 16 | 31 | 48 | 10 | 19 | 77 | 108 |
| **EQ-5D health state at 30 day follow-up** | 10 (7, 100) | 100 (60, 100) | 100 (89, 100) | 99 (70, 100) | 83 (9, 100) | 83 (70, 100) | 100 (95, 100) | 100 (45, 100) | 100 (50, 100) |
| Missing | 14 | 10 | 15 | 39 | 61 | 8 | 17 | 86 | 125 |
| **Cost of treatment** | 14,000 (500, 53,000) | 7,000 (0, 14,000) | 2,025 (500, 11,000) | 3,000 (500, 15,000) | 2,500 (500, 25,000) | 5,000 (0, 15,000) | 1,500 (200, 3,000) | 2,000 (299, 13,000) | 2,000 (500, 14,000) |
| Missing | 23 | 14 | 18 | 55 | 90 | 21 | 21 | 132 | 187 |

[1]Median (Q1, Q3); n (%)

### Results with 95% confidence intervals (CI)

#### Table S2. Outcomes in all patients during the entire study period, by treatment arm(95% CI)

| Outcome | ATLS, N = 44 | PTC, N = 130 | Standard care, N = 202 |
|---|---|---|---|
| **30 day mortality** | | | |
| Yes | 2.5 (0, 15) | 7.3 (2.9, 13) | 13 (7.49, 17) |
| No | 98 (85, 100) | 93 (86, 96) | 87 (81, 91) |
| Missing | 4 (1, 9) | 20 (12, 29) | 29 (19, 40) |
| **24 hour mortality** | | | |
| Yes | | | 0.5 (0, 1.6) |
| No | | | 99 (98, 99) |
| Missing | 4 (1, 9) | 13 (6, 20) | 7 (3, 12) |
| **In-hospital mortality** | | | |
| Yes | 2.3 (0, 13.39) | 5.4 (2.1, 10) | 11 (6.67, 15) |
| No | 98 (86, 100) | 95 (90, 98) | 90 (85, 93) |
| Missing | 1 (0, 3) | | 2 (0, 5) |
| **Self-ambulatory at discharge** | | | |
| Yes | 97 (81, 100) | 97 (91, 99) | 98 (93.35, 99) |
| No | 2.8 (0, 17.91) | 3.4 (0.9, 8.11) | 2.3 (0.6, 6) |
| Missing | 8 (3, 13.73) | 11 (5, 17) | 26 (16, 35) |
| **Return to work** | | | |
| Yes | 64 (40.57, 80) | 72 (62, 80) | 51 (42, 59) |
| No | 36 (18, 57) | 28 (18, 36) | 49 (39, 56) |
| Missing | 19 (11, 27) | 31 (21, 42) | 68 (53, 82) |
| **Pulmonary complication** | | | |
| Yes | | 0.9 (0, 3.5) | |
| No | | 99 (95, 100) | |
| Missing | 11 (5, 18) | 21 (13, 30) | 35 (24, 46) |
| **Septic complication** | | | |
| Yes | | 3.7 (0.9, 8.3) | 1.2 (0, 3.7) |
| No | | 96 (90, 98) | 99 (94, 99) |
| Missing | 11 (5, 18) | 22 (13, 31) | 34 (23, 45) |
| **Renal failure** | | | |
| Yes | | | |
| No | | | |
| Missing | 10 (4, 16) | 21 (13, 30) | 33 (21.57, 43) |
| **Coagulopathy** | | | |
| Yes | 3 (0, 16) | | |

| Outcome | ATLS, N = 44 | PTC, N = 130 | Standard care, N = 202 |
|---|---|---|---|
| No | 97 (84, 100) | | |
| Missing | 11 (5, 18) | 22 (14, 32) | 33 (21.57, 43) |
| **Need for reexploration or resurgery** | | | |
| Yes | | 1.8 (0, 5.6) | 1.9 (0, 5.16) |
| No | | 98 (93, 99) | 98 (94, 99) |
| Missing | 10 (5, 16.51) | 20 (12, 28) | 43 (31, 55) |
| **Failure of conservative management** | | | |
| No | 94 (79, 100) | 98 (92, 99) | 99 (95, 99) |
| Yes | 6.5 (0, 20) | 1.9 (0, 6.09) | 1.3 (0, 4.1) |
| Missing | 13 (7, 20) | 23 (14, 32) | 46 (33, 58) |
| **EQ-5D mobility at discharge** | | | |
| I have no problems in walking | 80 (58, 91) | 88 (77, 93) | 62 (54, 69) |
| I have some problems in walking | 10 (2.8, 27.13) | 7.9 (3.66, 15) | 18 (11, 23) |
| I am confined to bed | 10 (2.8, 25) | 4 (1.1, 9.95) | 20 (13, 26) |
| Missing | 14 (7.11, 22) | 29 (19, 39) | 45 (33, 57) |
| **EQ-5D self-care at discharge** | | | |
| I have no problems with self-care | 63 (42, 78.24) | 83 (74, 88) | 52 (43, 59) |
| I have some problems bathing or dressing myself | 13 (3.1, 27.28) | 9.9 (4.95, 16) | 27 (19.37, 33) |
| I am unable to bathe or dress myself | 23 (8.05, 39) | 6.9 (3, 13) | 21 (14, 27) |
| Missing | 14 (7.11, 22) | 29 (19, 39) | 45 (33, 57) |
| **EQ-5D usual activities at discharge** | | | |
| I have no problems in performing my usual activities | 57 (36.2, 74) | 81 (72, 87) | 48 (39, 54.34) |
| I have some problems in performing my usual activities | 23 (8.3, 40) | 11 (5.13, 17) | 36 (29, 43) |
| I am unable to perform my usual activities | 20 (6.75, 37) | 7.9 (3.7, 14.71) | 16 (9.88, 21) |
| Missing | 14 (7.11, 22) | 29 (19, 39) | 45 (33, 57) |
| **EQ-5D pain/discomfort at discharge** | | | |
| I have no pain or discomfort | 43 (26, 61.31) | 52 (41, 60) | 27 (20, 33) |
| I have moderate pain or discomfort | 57 (36.59, 72) | 44 (34, 53.16) | 36 (28, 43) |
| I have extreme pain or discomfort | | 4 (1, 9.38) | 36 (27.04, 43) |
| Missing | 14 (7.11, 22) | 29 (19, 39) | 45 (33, 57) |
| **EQ-5D anxiety/depression at discharge** | | | |
| I am not anxious or depressed | 70 (50, 85) | 86 (77.13, 91) | 89 (83, 93) |
| I am moderately anxious or depressed | 30 (13, 47) | 13 (6.92, 19) | 9.6 (5.84, 15) |
| I am extremely anxious or depressed | | 1 (0, 4.37) | 1.9 (0, 4.7) |
| Missing | 14 (7.11, 22) | 29 (19, 39) | 45 (33, 57) |
| **EQ-5D mobility at 30 day follow-up** | | | |
| I have no problems in walking | 77 (55.4, 89) | 95 (88, 97) | 74 (65, 80) |
| I have some problems in walking | 19 (6.39, 38) | 3 (0.9, 8.4) | 16 (9.25, 22) |

| Outcome | ATLS, N = 44 | PTC, N = 130 | Standard care, N = 202 |
|---|---|---|---|
| I am confined to bed | 3.8 (0, 23.3) | 2 (0, 6.18) | 9.8 (5.6, 16) |
| Missing | 18 (10, 26) | 29 (19, 39) | 69 (54, 83) |
| **EQ-5D self-care at 30 day follow-up** | | | |
| I have no problems with self-care | 81 (58, 92.79) | 94 (85.78, 97) | 77 (68, 82) |
| I have some problems bathing or dressing myself | 12 (0, 29.99) | 2 (0, 7.22) | 13 (7.2, 19) |
| I am unable to bathe or dress myself | 7.7 (0, 24.36) | 4 (1, 9.3) | 9.8 (5.7, 16) |
| Missing | 18 (10, 26) | 29 (19, 39) | 69 (54, 83) |
| **EQ-5D usual activities at 30 day follow-up** | | | |
| I have no problems in performing my usual activities | 81 (60, 93) | 93 (85, 96) | 79 (70, 85) |
| I have some problems in performing my usual activities | 12 (3.4, 31.69) | 3 (0.9, 8.5) | 14 (8.4, 20) |
| I am unable to perform my usual activities | 7.7 (0, 24.36) | 4 (1, 9.3) | 7.5 (3.8, 14) |
| Missing | 18 (10, 26) | 29 (19, 39) | 69 (54, 83) |
| **EQ-5D pain/discomfort at 30 day follow-up** | | | |
| I have no pain or discomfort | 77 (57, 91) | 82 (73, 88) | 78 (69, 84) |
| I have moderate pain or discomfort | 23 (7.7, 42) | 17 (9.3, 24) | 20 (13, 26) |
| I have extreme pain or discomfort | | 1 (0, 4.3) | 1.5 (0, 5.1) |
| Missing | 18 (10, 26) | 29 (19, 39) | 69 (54, 83) |
| **EQ-5D anxiety/depression at 30 day follow-up** | | | |
| I am not anxious or depressed | 88 (65, 96) | 92 (84, 95) | 95 (88, 97) |
| I am moderately anxious or depressed | 12 (3.4, 31.69) | 6.9 (2.9, 14) | 3.8 (1.5, 9) |
| I am extremely anxious or depressed | | 1 (0, 4.3) | 0.8 (0, 4.3) |
| Missing | 18 (10, 26) | 29 (19, 39) | 69 (54, 83) |
| **Patient satisfaction** | | | |
| Very satisfied | 74 (57, 86.02) | 85 (77, 90) | 69 (60, 74) |
| Somewhat satisfied | 21 (8.8, 34) | 9.9 (5, 16) | 16 (10, 20) |
| Somewhat dissatisfied | 2.6 (0, 13) | 1.7 (0, 5.15) | 9 (5.1, 14) |
| Very dissatisfied | 2.6 (0, 12.43) | 3.3 (0.8, 7.71) | 5.1 (2.3, 9.06) |
| Missing | 6 (2, 11) | 9 (3, 15) | 25 (15, 34) |
| **Number of hospitalizations for this injury** | | | |
| Missing | 13 (7, 20) | 19 (11, 27) | 36 (25, 47) |
| **EQ-5D health state at discharge** | 60 (50, 60) | 80 (79, 79) | 50 (4, 50) |
| Missing | 15 (8, 23) | 35 (25, 47) | 58 (44, 72) |
| **EQ-5D health state at 30 day follow-up** | 88 (80, 100) | 100 (99, 99) | 80 (9, 90) |
| Missing | 18 (10, 26) | 32 (22, 44) | 75 (59, 90) |
| **Cost of treatment** | 5000 (0, 14000) | 2000 (1040.14, 2499) | 3000 (1000, 8000) |
| Missing | 35 (24, 48) | 39 (29, 51.46) | 113 (96, 130) |

#### Table S3. Outcomes in all patients before training, by treatment arm(95% CI)

| Outcome | ATLS, N = 16 | PTC, N = 57 | Standard care, N = 41 |
|---|---|---|---|
| **30 day mortality** | | | |
| Yes | | 10 (2.32, 20) | 2.6 (0, 14.5) |
| No | | 90 (78.57, 96) | 97 (81, 100) |
| Missing | 2 (0, 5) | 8 (3, 14) | 3 (0, 7) |
| **24 hour mortality** | | | |
| Yes | | | 2.4 (0, 11.05) |
| No | | | 98 (88, 100) |
| Missing | 2 (0, 5) | 5 (1, 9) | |
| **In-hospital mortality** | | | |
| Yes | | 7 (1.9, 16) | 4.9 (0, 17) |
| No | | 93 (81.7, 97) | 95 (81, 98) |
| Missing | | | |
| **Self-ambulatory at discharge** | | | |
| Yes | 92 (55.29, 100) | 96 (84, 98) | 95 (78.8, 100) |
| No | 8.3 (0, 42.23) | 4.1 (0, 13.82) | 5.1 (0, 17) |
| Missing | 4 (1, 8) | 8 (3, 13.13) | 2 (0, 5) |
| **Return to work** | | | |
| Yes | 83 (0, 100) | 74 (58, 85) | 41 (23, 58) |
| No | 17 (0, 70.86) | 26 (12, 39) | 59 (37.73, 75) |
| Missing | 10 (4, 16) | 15 (8, 22) | 12 (6, 19) |
| **Pulmonary complication** | | | |
| Yes | | 2.2 (0, 11.73) | |
| No | | 98 (88, 100) | |
| Missing | 6 (2, 11) | 11 (5, 18) | 2 (0, 5) |
| **Septic complication** | | | |
| Yes | | 4.4 (0, 14) | 2.5 (0, 16.45) |
| No | | 96 (85.26, 100) | 98 (83, 100) |
| Missing | 6 (2, 11) | 12 (6, 19) | 1 (0, 3) |
| **Renal failure** | | | |
| Yes | | | |
| No | | | |
| Missing | 5 (1, 10) | 11 (5, 18) | 1 (0, 3) |
| **Coagulopathy** | | | |
| Yes | | | |
| No | | | |
| Missing | 6 (2, 11) | 12 (6, 19) | 1 (0, 3) |
| **Need for reexploration or resurgery** | | | |

| Outcome | ATLS, N = 16 | PTC, N = 57 | Standard care, N = 41 |
|---|---|---|---|
| Yes | | 2.1 (0, 11.23) | |
| No | | 98 (86, 100) | |
| Missing | 4 (1, 8) | 10 (4, 16) | 1 (0, 3) |
| **Failure of conservative management** | | | |
| No | | 98 (85, 100) | |
| Yes | | 2.2 (0, 14.13) | |
| Missing | 10 (4, 16) | 11 (5, 17) | 2 (0, 5) |
| **EQ-5D mobility at discharge** | | | |
| I have no problems in walking | 67 (30.53, 89) | 83 (66.74, 92) | 62 (44, 75) |
| I have some problems in walking | 8.3 (0, 36.88) | 12 (2.6, 24) | 18 (7.1, 31.39) |
| I am confined to bed | 25 (0, 56) | 4.9 (0, 16.18) | 21 (9.3, 35) |
| Missing | 4 (1, 8) | 16 (8, 24) | 2 (0, 5) |
| **EQ-5D self-care at discharge** | | | |
| I have no problems with self-care | 58 (25, 87.2) | 80 (64, 90) | 59 (41, 73.74) |
| I have some problems bathing or dressing myself | 17 (0, 50) | 12 (2.97, 24) | 15 (4.97, 27.42) |
| I am unable to bathe or dress myself | 25 (0, 56) | 7.3 (2.2, 20) | 26 (13, 40) |
| Missing | 4 (1, 8) | 16 (8, 24) | 2 (0, 5) |
| **EQ-5D usual activities at discharge** | | | |
| I have no problems in performing my usual activities | 50 (17, 75) | 76 (61, 87) | 54 (38, 69) |
| I have some problems in performing my usual activities | 25 (0, 54.17) | 15 (5.7, 28) | 28 (14, 42) |
| I am unable to perform my usual activities | 25 (0, 56) | 9.8 (2.7, 23) | 18 (7.1, 32) |
| Missing | 4 (1, 8) | 16 (8, 24) | 2 (0, 5) |
| **EQ-5D pain/discomfort at discharge** | | | |
| I have no pain or discomfort | 33 (8.3, 63) | 34 (20, 50) | 33 (19, 48.33) |
| I have moderate pain or discomfort | 67 (30.09, 89) | 63 (44.05, 76) | 41 (24, 56) |
| I have extreme pain or discomfort | | 2.4 (0, 13) | 26 (14, 40.01) |
| Missing | 4 (1, 8) | 16 (8, 24) | 2 (0, 5) |
| **EQ-5D anxiety/depression at discharge** | | | |
| I am not anxious or depressed | 50 (17, 75) | 73 (57, 85) | 79 (62.02, 89) |
| I am moderately anxious or depressed | 50 (14, 75) | 24 (11, 38) | 18 (6.97, 32) |
| I am extremely anxious or depressed | | 2.4 (0, 13) | 2.6 (0, 14) |
| Missing | 4 (1, 8) | 16 (8, 24) | 2 (0, 5) |
| **EQ-5D mobility at 30 day follow-up** | | | |
| I have no problems in walking | 67 (0, 100) | 91 (77.53, 97) | 65 (46.25, 81) |
| I have some problems in walking | 17 (0, 70.43) | 7 (2.02, 19) | 13 (3.12, 27) |
| I am confined to bed | 17 (0, 70.86) | 2.3 (0, 13.05) | 23 (9.02, 39) |
| Missing | 10 (4, 16) | 14 (7, 21) | 10 (4, 17) |
| **EQ-5D self-care at 30 day follow-up** | | | |
| I have no problems with self-care | 83 (0.05, 100) | 91 (79.35, 97) | 71 (53, 85) |

| Outcome | ATLS, N = 16 | PTC, N = 57 | Standard care, N = 41 |
|---|---|---|---|
| I have some problems bathing or dressing myself | 17 (0, 70.43) | 4.7 (0, 16) | 6.5 (0, 31.44) |
| I am unable to bathe or dress myself | | 4.7 (0, 15.32) | 23 (11, 40) |
| Missing | 10 (4, 16) | 14 (7, 21) | 10 (4, 17) |
| **EQ-5D usual activities at 30 day follow-up** | | | |
| I have no problems in performing my usual activities | | 91 (79.35, 97) | 71 (52.15, 85) |
| I have some problems in performing my usual activities | | 4.7 (0, 16) | 19 (6.7, 35) |
| I am unable to perform my usual activities | | 4.7 (0, 15.32) | 9.7 (2.46, 25.76) |
| Missing | 10 (4, 16) | 14 (7, 21) | 10 (4, 17) |
| **EQ-5D pain/discomfort at 30 day follow-up** | | | |
| I have no pain or discomfort | 67 (0, 100) | 77 (61.95, 88) | 74 (52.26, 85) |
| I have moderate pain or discomfort | 33 (0, 75) | 21 (8.6, 33) | 26 (12, 44.69) |
| I have extreme pain or discomfort | | 2.3 (0, 12.93) | |
| Missing | 10 (4, 16) | 14 (7, 21) | 10 (4, 17) |
| **EQ-5D anxiety/depression at 30 day follow-up** | | | |
| I am not anxious or depressed | | 86 (71.41, 94) | 97 (81, 100) |
| I am moderately anxious or depressed | | 12 (2.9, 24.57) | 3.2 (0, 18.59) |
| I am extremely anxious or depressed | | 2.3 (0, 12.93) | |
| Missing | 10 (4, 16) | 14 (7, 21) | 10 (4, 17) |
| **Patient satisfaction** | | | |
| Very satisfied | 64 (33.23, 87) | 73 (60, 83) | 67 (49, 79) |
| Somewhat satisfied | 36 (11, 63) | 16 (6.66, 27) | 23 (11, 35.02) |
| Somewhat dissatisfied | | 3.9 (0, 13) | 7.7 (2.2, 21) |
| Very dissatisfied | | 7.8 (2.1, 18.06) | 2.6 (0, 17) |
| Missing | 2 (0, 5) | 6 (2, 11) | 2 (0, 5) |
| **Number of hospitalizations for this injury** | | | |
| Missing | 10 (4, 16) | 8 (3, 14) | 2 (0, 5) |
| **EQ-5D health state at discharge** | 60 (40, 81.8) | 80 (60, 80) | 50 (3, 80) |
| Missing | 5 (1, 10) | 16 (8, 24) | 10 (4, 17) |
| **EQ-5D health state at 30 day follow-up** | 100 (55, 100) | 100 (90, 100) | 10 (8, 100) |
| Missing | 10 (4, 16) | 15 (8.11, 23) | 14 (8, 22) |
| **Cost of treatment** | 7000 (0, 14000) | 2025 (600, 6000) | 14000 (500, 35000) |
| Missing | 14 (7, 22) | 18 (11, 26) | 23 (14.12, 33) |

#### Table S4. Outcomes in all patients after training, by treatment arm(95% CI)

| Outcome | ATLS, N = 28 | PTC, N = 73 | Standard care, N = 161 |
|---|---|---|---|
| **30 day mortality** | | | |
| Yes | 3.8 (0, 20.98) | 4.9 (0, 13) | 16 (9.28, 21) |
| No | 96 (78, 100) | 95 (84, 98) | 84 (77, 89) |
| Missing | 2 (0, 5) | 12 (6, 19) | 26 (17, 36) |
| **24 hour mortality** | | | |
| Yes | | | |
| No | | | |
| Missing | 2 (0, 5) | 8 (3, 14) | 7 (3, 12) |
| **In-hospital mortality** | | | |
| Yes | 3.7 (0, 20.68) | 4.1 (0, 11) | 12 (6.43, 16) |
| No | 96 (79, 100) | 96 (87.42, 99) | 88 (81, 91) |
| Missing | 1 (0, 3) | | 2 (0, 5) |
| **Self-ambulatory at discharge** | | | |
| Yes | | 97 (86, 99) | 99 (94, 100) |
| No | | 2.9 (0, 9.08) | 1.5 (0, 5.59) |
| Missing | 4 (1, 8) | 3 (0, 7) | 24 (14, 32.64) |
| **Return to work** | | | |
| Yes | 58 (33, 78) | 70 (55, 80) | 53 (43, 61.06) |
| No | 42 (21, 66.95) | 30 (18, 42.29) | 47 (36, 56) |
| Missing | 9 (4, 15) | 16 (9, 24) | 56 (43, 69.34) |
| **Pulmonary complication** | | | |
| Yes | | | |
| No | | | |
| Missing | 5 (1, 9) | 10 (5, 17) | 33 (22, 43) |
| **Septic complication** | | | |
| Yes | | 3.2 (0, 11) | 0.8 (0, 4) |
| No | | 97 (88, 100) | 99 (96, 99) |
| Missing | 5 (1, 9) | 10 (5, 17) | 33 (22, 43) |
| **Renal failure** | | | |
| Yes | | | |
| No | | | |
| Missing | 5 (1, 9) | 10 (5, 17) | 32 (21, 42) |
| **Coagulopathy** | | | |
| Yes | 4.3 (0, 22.71) | | |
| No | 96 (77, 100) | | |
| Missing | 5 (1, 9) | 10 (5, 17) | 32 (21, 42) |
| **Need for reexploration or resurgery** | | | |

| Outcome | ATLS, N = 28 | PTC, N = 73 | Standard care, N = 161 |
|---|---|---|---|
| Yes | | 1.6 (0, 9.21) | 2.5 (0, 7) |
| No | | 98 (90, 100) | 97 (92, 98) |
| Missing | 6 (2, 11) | 10 (5, 17) | 42 (30.18, 54) |
| **Failure of conservative management** | | | |
| No | 92 (71, 100) | 98 (90, 100) | 98 (93, 99) |
| Yes | 8 (0, 24.44) | 1.6 (0, 7.99) | 1.7 (0, 5.8) |
| Missing | 3 (0, 6) | 12 (6, 19.14) | 44 (32, 56) |
| **EQ-5D mobility at discharge** | | | |
| I have no problems in walking | 89 (60, 100) | 92 (80.1, 97) | 62 (52, 70) |
| I have some problems in walking | 11 (0, 33) | 5 (1.5, 13.83) | 19 (12, 27) |
| I am confined to bed | | 3.3 (0, 11) | 19 (11, 26) |
| Missing | 10 (4, 17) | 13 (6, 21) | 43 (30.04, 54) |
| **EQ-5D self-care at discharge** | | | |
| I have no problems with self-care | 67 (34.68, 87) | 85 (72, 92) | 50 (40, 58) |
| I have some problems bathing or dressing myself | 11 (0, 33.5) | 8.3 (2.56, 17.71) | 31 (22, 39) |
| I am unable to bathe or dress myself | 22 (4.52, 44) | 6.7 (1.76, 16) | 19 (11, 25) |
| Missing | 10 (4, 17) | 13 (6, 21) | 43 (30.04, 54) |
| **EQ-5D usual activities at discharge** | | | |
| I have no problems in performing my usual activities | 61 (33, 82.01) | 85 (72, 92) | 47 (38, 56) |
| I have some problems in performing my usual activities | 22 (5.6, 50) | 8.3 (2.56, 17.71) | 38 (28, 45) |
| I am unable to perform my usual activities | 17 (0, 39.02) | 6.7 (1.76, 16) | 15 (8.29, 21) |
| Missing | 10 (4, 17) | 13 (6, 21) | 43 (30.04, 54) |
| **EQ-5D pain/discomfort at discharge** | | | |
| I have no pain or discomfort | 50 (25, 71) | 65 (52, 76) | 25 (16.01, 32) |
| I have moderate pain or discomfort | 50 (22, 69) | 30 (18, 41) | 35 (27, 44) |
| I have extreme pain or discomfort | | 5 (1.5, 14) | 40 (31, 48) |
| Missing | 10 (4, 17) | 13 (6, 21) | 43 (30.04, 54) |
| **EQ-5D anxiety/depression at discharge** | | | |
| I am not anxious or depressed | 83 (57, 95.75) | 95 (84.84, 98) | 92 (85, 96) |
| I am moderately anxious or depressed | 17 (0, 41) | 5 (0.6, 13) | 6.8 (3.4, 13) |
| I am extremely anxious or depressed | | | 1.7 (0, 5.3) |
| Missing | 10 (4, 17) | 13 (6, 21) | 43 (30.04, 54) |
| **EQ-5D mobility at 30 day follow-up** | | | |
| I have no problems in walking | 80 (53, 93.32) | 98 (90, 100) | 77 (67, 83.81) |
| I have some problems in walking | 20 (5.9, 44) | | 17 (9.4, 24) |
| I am confined to bed | | 1.7 (0, 8.79) | 5.9 (2.1, 12) |
| Missing | 8 (3, 14) | 15 (8, 23) | 59 (46, 72.14) |
| **EQ-5D self-care at 30 day follow-up** | | | |
| I have no problems with self-care | 80 (56, 94) | 97 (88, 100) | 79 (68.15, 85) |

| Outcome | ATLS, N = 28 | PTC, N = 73 | Standard care, N = 161 |
| --- | --- | --- | --- |
| I have some problems bathing or dressing myself | 10 (0, 31) | | 15 (8.23, 23) |
| I am unable to bathe or dress myself | 10 (0, 31.74) | 3.4 (0, 12) | 5.9 (2.13, 12) |
| Missing | 8 (3, 14) | 15 (8, 23) | 59 (46, 72.14) |
| **EQ-5D usual activities at 30 day follow-up** | | | |
| I have no problems in performing my usual activities | 75 (50, 91) | 95 (83.7, 98) | 81 (70, 87) |
| I have some problems in performing my usual activities | 15 (4.2, 38) | 1.7 (0, 7.68) | 12 (6.3, 19) |
| I am unable to perform my usual activities | 10 (0, 31.74) | 3.4 (0, 12) | 6.9 (2.9, 13) |
| Missing | 8 (3, 14) | 15 (8, 23) | 59 (46, 72.14) |
| **EQ-5D pain/discomfort at 30 day follow-up** | | | |
| I have no pain or discomfort | 80 (56.42, 94) | 86 (73, 93) | 79 (69, 85) |
| I have moderate pain or discomfort | 20 (5.3, 41.4) | 14 (5.3, 23.93) | 19 (11, 27) |
| I have extreme pain or discomfort | | | 2 (0, 8.07) |
| Missing | 8 (3, 14) | 15 (8, 23) | 59 (46, 72.14) |
| **EQ-5D anxiety/depression at 30 day follow-up** | | | |
| I am not anxious or depressed | 85 (60.19, 95) | 97 (88.33, 100) | 95 (87, 97) |
| I am moderately anxious or depressed | 15 (4.2, 38) | 3.4 (0, 11) | 3.9 (1, 9.9) |
| I am extremely anxious or depressed | | | 1 (0, 4.95) |
| Missing | 8 (3, 14) | 15 (8, 23) | 59 (46, 72.14) |
| **Patient satisfaction** | | | |
| Very satisfied | 79 (57, 93) | 94 (83, 97) | 70 (61, 76) |
| Somewhat satisfied | 13 (3.29, 32) | 5.7 (1.5, 13) | 14 (8.04, 19) |
| Somewhat dissatisfied | 4.2 (0, 20.59) | | 9.4 (5.58, 16) |
| Very dissatisfied | 4.2 (0, 20.74) | | 5.8 (2.2, 10) |
| Missing | 4 (1, 8) | 3 (0, 7) | 23 (13, 31) |
| **Number of hospitalizations for this injury** | | | |
| Missing | 3 (0, 6) | 11 (5, 18) | 34 (23.34, 46) |
| **EQ-5D health state at discharge** | 65 (50, 80) | 90 (80, 90) | 50 (4, 50) |
| Missing | 10 (4, 17) | 19 (11, 28) | 48 (35, 61) |
| **EQ-5D health state at 30 day follow-up** | 83 (70, 100) | | 83 (10, 100) |
| Missing | 8 (3, 14) | 17 (9, 25.63) | 61 (47, 74) |
| **Cost of treatment** | 5000 (0, 15000) | 1500 (250, 1600) | 2500 (1000, 8000) |
| Missing | 21 (12, 30) | 21 (12.87, 30) | 90 (74, 106) |

#### Table S5. Absolute change from baseline for all outcomes, comparing the period after training with the period before training, by treatment arms(95% CI)

| Outcome | ATLS, N = 28 | PTC, N = 73 | Standard care, N = 161 |
|---|---|---|---|
| **30 day mortality** | | | |
| Yes | 3.8 (0, 20.98) | -5.1 (-16.57, 4.76) | 13.4 (3, 20) |
| No | -4 (-22, 0) | 5 (-6, 14) | -13 (-20, -3) |
| Missing | 0 (-4.9, 3) | 4 (-5, 12) | 23 (14, 34) |
| **24 hour mortality** | | | |
| Yes | | | -2.4 (-12, 0) |
| No | | | 2 (0, 7) |
| Missing | 0 (-4.9, 3) | 3 (-4, 10) | 7 (3, 12) |
| **In-hospital mortality** | | | |
| Yes | 3.7 (0, 20.68) | -2.9 (-12.79, 4.2) | 7.1 (-4, 13.3) |
| No | -4 (-21, 0) | 3 (-5, 11) | -7 (-14, 2) |
| Missing | 1 (0, 3) | | 2 (0, 5) |
| **Self-ambulatory at discharge** | | | |
| Yes | 8 (0, 37) | 1 (-6, 8) | 4 (-2, 14) |
| No | -8.3 (-44.67, 0) | -1.2 (-9.5, 4.8) | -3.6 (-14.56, 1.5) |
| Missing | 0 (-7, 4) | -5 (-13.26, 0) | 22 (12, 30) |
| **Return to work** | | | |
| Yes | -25 (-56, 21) | -4 (-24, 12) | 12 (-10, 30) |
| No | 25 (-27.41, 55) | 4 (-15, 21) | -12 (-35, 7) |
| Missing | -1 (-11, 6) | 1 (-10.1, 11) | 44 (29, 58) |
| **Pulmonary complication** | | | |
| Yes | | -2.2 (-12, 0) | |
| No | | 2 (0, 7) | |
| Missing | -1 (-9, 4) | -1 (-11, 7) | 31 (19, 41) |
| **Septic complication** | | | |
| Yes | | -1.2 (-10.31, 5.5) | -1.7 (-11.72, 1.6) |
| No | | 1 (-6, 8.2) | 1 (-2, 7) |
| Missing | -1 (-9, 4) | -2 (-12, 6) | 32 (21, 42) |
| **Renal failure** | | | |
| Yes | | | |
| No | | | |
| Missing | 0 (-7, 5) | -1 (-11, 7) | 31 (20, 41) |
| **Coagulopathy** | | | |
| Yes | 4.3 (0, 22.71) | | |
| No | -4 (-23, 0) | | |

| Outcome | ATLS, N = 28 | PTC, N = 73 | Standard care, N = 161 |
|---|---|---|---|
| Missing | -1 (-9, 4) | -2 (-12, 5) | 31 (20, 41) |
| **Need for reexploration or resurgery** | | | |
| Yes | | -0.5 (-8.44, 3.3) | 2.5 (0, 7) |
| No | | 0 (-5.44, 4) | -3 (-8, -2) |
| Missing | 2 (-5, 7) | 0 (-9, 8) | 41 (29, 54) |
| **Failure of conservative management** | | | |
| No | -8 (-29, 0) | 0 (-6.06, 4) | -2 (-7, -1) |
| Yes | 8 (0, 24.45) | -0.6 (-9.45, 3.2) | 1.7 (0, 5.8) |
| Missing | -7 (-16.47, -2) | 1 (-8, 10) | 42 (29, 54) |
| **EQ-5D mobility at discharge** | | | |
| I have no problems in walking | 22 (-12, 56) | 9 (-5, 23) | 0 (-17, 19.83) |
| I have some problems in walking | 2.7 (-23.17, 20) | -7 (-19.12, 4.99) | 1 (-16.13, 14) |
| I am confined to bed | -25 (-60.74, -7.1) | -1.6 (-11.5, 5.8) | -2 (-22, 10) |
| Missing | 6 (-2, 12) | -3 (-14, 7) | 41 (29, 53) |
| **EQ-5D self-care at discharge** | | | |
| I have no problems with self-care | 9 (-31.87, 47.44) | 5 (-11, 20) | -9 (-27, 8) |
| I have some problems bathing or dressing myself | -6 (-39.79, 15) | -3.7 (-16.9, 8.16) | 16 (0, 29.13) |
| I am unable to bathe or dress myself | -3 (-42.58, 26.36) | -0.6 (-11.51, 10) | -7 (-25.46, 6) |
| Missing | 6 (-2, 12) | -3 (-14, 7) | 41 (29, 53) |
| **EQ-5D usual activities at discharge** | | | |
| I have no problems in performing my usual activities | 11 (-30.58, 46) | 9 (-9, 23.11) | -7 (-26, 11) |
| I have some problems in performing my usual activities | -3 (-37, 28.8) | -6.7 (-21.53, 4.5) | 10 (-7.49, 26) |
| I am unable to perform my usual activities | -8 (-43, 21) | -3.1 (-15.5, 7.7) | -3 (-20, 8.18) |
| Missing | 6 (-2, 12) | -3 (-14, 7) | 41 (29, 53) |
| **EQ-5D pain/discomfort at discharge** | | | |
| I have no pain or discomfort | 17 (-23, 51) | 31 (9, 47) | -8 (-26.97, 6) |
| I have moderate pain or discomfort | -17 (-52, 20) | -33 (-52.63, -12) | -6 (-24, 13) |
| I have extreme pain or discomfort | | 2.6 (-4.7, 9.7) | 14 (-6.34, 27) |
| Missing | 6 (-2, 12) | -3 (-14, 7) | 41 (29, 53) |
| **EQ-5D anxiety/depression at discharge** | | | |
| I am not anxious or depressed | 33 (-5.81, 64) | 22 (7, 37) | 13 (0, 29) |
| I am moderately anxious or depressed | -33 (-67, 3) | -19 (-35.39, -5.8) | -11.2 (-27.48, 0.4) |
| I am extremely anxious or depressed | | -2.4 (-17, 0) | -0.9 (-10.47, 2.5) |
| Missing | 6 (-2, 12) | -3 (-14, 7) | 41 (29, 53) |
| **EQ-5D mobility at 30 day follow-up** | | | |
| I have no problems in walking | 13 (-29.15, 61) | 7 (-2, 17) | 12 (-9, 30) |
| I have some problems in walking | 3 (-50, 29) | -7 (-19.34, -2.1) | 4 (-14, 15) |

| Outcome | ATLS, N = 28 | PTC, N = 73 | Standard care, N = 161 |
|---|---|---|---|
| I am confined to bed | -17 (-100, 0) | -0.6 (-8.96, 3.82) | -17.1 (-36.1, -3.84) |
| Missing | -2 (-12, 5) | 1 (-10, 11) | 49 (34, 63) |
| **EQ-5D self-care at 30 day follow-up** | | | |
| I have no problems with self-care | -3 (-28.6, 53) | 6 (-4, 16) | 8 (-11, 26) |
| I have some problems bathing or dressing myself | -7 (-64.7, 16.07) | -4.7 (-17.2, 0) | 8.5 (-5, 18) |
| I am unable to bathe or dress myself | 10 (0, 31.59) | -1.3 (-10.45, 5.7) | -17.1 (-35.9, -4.9) |
| Missing | -2 (-12, 5) | 1 (-10, 11) | 49 (34, 63) |
| **EQ-5D usual activities at 30 day follow-up** | | | |
| I have no problems in performing my usual activities | -25 (-50, -9) | 4 (-8, 13) | 10 (-8.1, 28.5) |
| I have some problems in performing my usual activities | 15 (4.2, 38) | -3 (-14, 2.1) | -7 (-24.6, 6) |
| I am unable to perform my usual activities | 10 (0, 31.59) | -1.3 (-10.45, 5.7) | -2.8 (-17.58, 6.4) |
| Missing | -2 (-12, 5) | 1 (-10, 11) | 49 (34, 63) |
| **EQ-5D pain/discomfort at 30 day follow-up** | | | |
| I have no pain or discomfort | 13 (-30, 62) | 9 (-8, 23) | 5 (-10, 23) |
| I have moderate pain or discomfort | -13 (-65, 28) | -7 (-23.4, 8) | -7 (-27, 7) |
| I have extreme pain or discomfort | | -2.3 (-16, 0) | 2 (0, 8.07) |
| Missing | -2 (-12, 5) | 1 (-10, 11) | 49 (34, 63) |
| **EQ-5D anxiety/depression at 30 day follow-up** | | | |
| I am not anxious or depressed | -15 (-39.72, -5) | 11 (0, 24) | -2 (-8, 8) |
| I am moderately anxious or depressed | 15 (4.2, 38) | -8.6 (-22, 1.02) | 0.7 (-13.46, 5.3) |
| I am extremely anxious or depressed | | -2.3 (-16, 0) | 1 (0, 4.95) |
| Missing | -2 (-12, 5) | 1 (-10, 11) | 49 (34, 63) |
| **Patient satisfaction** | | | |
| Very satisfied | 15 (-15, 45) | 21 (8, 32) | 3 (-14, 19) |
| Somewhat satisfied | -23 (-54, 4) | -10.3 (-22.87, 0) | -9 (-25.8, 3) |
| Somewhat dissatisfied | 4.2 (0, 20.59) | -3.9 (-13, 0) | 1.7 (-11.7, 9.2) |
| Very dissatisfied | 4.2 (0, 20.74) | -7.8 (-20, -2.28) | 3.2 (-8, 7.6) |
| Missing | 2 (-3, 6) | -3 (-10, 2) | 21 (11, 30) |
| **Number of hospitalizations for this injury** | 0 (-1, 0) | | |
| Missing | -7 (-16.47, -2) | 3 (-6, 12) | 32 (21, 44) |
| **EQ-5D health state at discharge** | 5 (-30, 30) | 10 (-10, 10) | 0 (-68.09, 55) |
| Missing | 5 (-3.48, 11) | 3 (-9, 14) | 38 (22, 52) |
| **EQ-5D health state at 30 day follow-up** | -17 (-33.16, 20) | | 73 (-13, 92) |
| Missing | -2 (-12, 5) | 2 (-10, 12) | 47 (30, 61.57) |
| **Cost of treatment** | -2000 (-14000, 13000) | -525 (-4922.65, 1000) | -11500 (-49000, 2250) |
| Missing | 7 (-7, 17) | 3 (-10, 14) | 67 (45.64, 86) |

#### Table S6. Relative change from baseline for all outcomes, comparing the period after training with the period before training, by treatment arms(95% CI)

| Outcome | ATLS, N = 28 | PTC, N = 73 | Standard care, N = 161 |
|---|---|---|---|
| **30 day mortality** | | | |
| Yes | | 0.49 (0, 2.31) | 6.15 (2.62, 10.91) |
| No | 0.96 (0.78, 1) | 1.06 (0.95, 1.2) | 0.87 (0.8, 0.97) |
| Missing | 1 (0, 4) | 1.5 (0.56, 4) | 8.67 (2.89, 27.17) |
| **24 hour mortality** | | | |
| Yes | | | |
| No | | | 1.02 (1, 1.08) |
| Missing | 1 (0, 4) | 1.6 (0.5, 6) | |
| **In-hospital mortality** | | | |
| Yes | | 0.59 (0, 2.89) | 2.45 (0.77, 6.19) |
| No | 0.96 (0.79, 1) | 1.03 (0.95, 1.15) | 0.93 (0.86, 1.03) |
| Missing | | | |
| **Self-ambulatory at discharge** | | | |
| Yes | 1.09 (1, 1.59) | 1.01 (0.95, 1.1) | 1.04 (0.98, 1.17) |
| No | | 0.71 (0, 4) | 0.29 (0, 1.52) |
| Missing | 1 (0, 3.5) | 0.38 (0, 1.57) | 12 (3.73, 29) |
| **Return to work** | | | |
| Yes | 0.7 (0.41, 1.42) | 0.95 (0.73, 1.2) | 1.29 (0.85, 2.24) |
| No | 2.47 (1.11, 7.14) | 1.15 (0.61, 2.48) | 0.8 (0.55, 1.2) |
| Missing | 0.9 (0.29, 2.25) | 1.07 (0.5, 2.25) | 4.67 (2.52, 9) |
| **Pulmonary complication** | | | |
| Yes | | | |
| No | | 1.02 (1, 1.08) | |
| Missing | 0.83 (0.14, 2.67) | 0.91 (0.36, 2.5) | 16.5 (5.11, 39) |
| **Septic complication** | | | |
| Yes | | 0.73 (0, 3.81) | 0.32 (0, 1.62) |
| No | | 1.01 (0.94, 1.1) | 1.01 (0.98, 1.08) |
| Missing | 0.83 (0.14, 2.67) | 0.83 (0.33, 2) | 33 (12.33, 44.32) |
| **Renal failure** | | | |
| Yes | | | |
| No | | | |
| Missing | 1 (0.11, 3) | 0.91 (0.36, 2.5) | 32 (11.5, 42) |
| **Coagulopathy** | | | |
| Yes | | | |
| No | 0.96 (0.77, 1) | | |
| Missing | 0.83 (0.14, 2.67) | 0.83 (0.33, 2.07) | 32 (11.5, 42) |

| Outcome | ATLS, N = 28 | PTC, N = 73 | Standard care, N = 161 |
|---|---|---|---|
| **Need for reexploration or resurgery** | | | |
| Yes | | 0.76 (0, 4.47) | |
| No | | 1 (0.95, 1.04) | 0.97 (0.92, 0.98) |
| Missing | 1.5 (0.26, 6) | 1 (0.36, 2.5) | 42 (16.32, 55.43) |
| **Failure of conservative management** | | | |
| No | 0.92 (0.71, 1) | 1 (0.94, 1.04) | 0.98 (0.93, 0.99) |
| Yes | | 0.73 (0, 4.65) | |
| Missing | 0.3 (0, 1.21) | 1.09 (0.47, 2.6) | 22 (6.82, 51) |
| **EQ-5D mobility at discharge** | | | |
| I have no problems in walking | 1.33 (0.88, 2.47) | 1.11 (0.96, 1.33) | 1 (0.77, 1.42) |
| I have some problems in walking | 1.33 (0.27, 5.5) | 0.42 (0, 2.49) | 1.06 (0.48, 3.01) |
| I am confined to bed | | 0.67 (0, 4.06) | 0.9 (0.42, 1.87) |
| Missing | 2.5 (0.62, 8) | 0.81 (0.37, 1.85) | 21.5 (7.3, 50) |
| **EQ-5D self-care at discharge** | | | |
| I have no problems with self-care | 1.16 (0.63, 2.62) | 1.06 (0.88, 1.31) | 0.85 (0.62, 1.22) |
| I have some problems bathing or dressing myself | 0.65 (0, 3.36) | 0.69 (0.14, 2.77) | 2.07 (1, 6.19) |
| I am unable to bathe or dress myself | 0.88 (0.09, 3.72) | 0.92 (0.18, 4.6) | 0.73 (0.37, 1.38) |
| Missing | 2.5 (0.62, 8) | 0.81 (0.37, 1.85) | 21.5 (7.3, 50) |
| **EQ-5D usual activities at discharge** | | | |
| I have no problems in performing my usual activities | 1.22 (0.58, 2.89) | 1.12 (0.93, 1.37) | 0.87 (0.63, 1.31) |
| I have some problems in performing my usual activities | 0.88 (0.17, 3.75) | 0.55 (0.11, 1.75) | 1.36 (0.86, 2.72) |
| I am unable to perform my usual activities | 0.68 (0, 3.3) | 0.68 (0.12, 3.59) | 0.83 (0.38, 2.01) |
| Missing | 2.5 (0.62, 8) | 0.81 (0.37, 1.85) | 21.5 (7.3, 50) |
| **EQ-5D pain/discomfort at discharge** | | | |
| I have no pain or discomfort | 1.52 (0.6, 4.73) | 1.91 (1.2, 3.22) | 0.76 (0.45, 1.36) |
| I have moderate pain or discomfort | 0.75 (0.35, 1.49) | 0.48 (0.28, 0.77) | 0.85 (0.57, 1.54) |
| I have extreme pain or discomfort | | 2.08 (0.65, 6.54) | 1.54 (0.87, 2.73) |
| Missing | 2.5 (0.62, 8) | 0.81 (0.37, 1.85) | 21.5 (7.3, 50) |
| **EQ-5D anxiety/depression at discharge** | | | |
| I am not anxious or depressed | 1.66 (0.94, 3.8) | 1.3 (1.1, 1.65) | 1.16 (1.01, 1.51) |
| I am moderately anxious or depressed | 0.34 (0, 1.29) | 0.21 (0, 0.75) | 0.38 (0.13, 1.23) |
| I am extremely anxious or depressed | | | 0.65 (0.14, 2.38) |
| Missing | 2.5 (0.62, 8) | 0.81 (0.37, 1.85) | 21.5 (7.3, 50) |
| **EQ-5D mobility at 30 day follow-up** | | | |
| I have no problems in walking | 1.19 (0.7, 3.16) | 1.08 (0.99, 1.22) | 1.18 (0.9, 1.62) |
| I have some problems in walking | 1.18 (0.42, 4) | | 1.31 (0.5, 4.52) |
| I am confined to bed | | 0.74 (0, 4.24) | 0.26 (0.07, 0.87) |
| Missing | 0.8 (0.23, 2) | 1.07 (0.5, 2.25) | 5.9 (3.16, 12.8) |
| **EQ-5D self-care at 30 day follow-up** | | | |

| Outcome | ATLS, N = 28 | PTC, N = 73 | Standard care, N = 161 |
|---|---|---|---|
| I have no problems with self-care | 0.96 (0.71, 2.58) | 1.07 (0.97, 1.21) | 1.11 (0.89, 1.47) |
| I have some problems bathing or dressing myself | 0.59 (0.12, 3.12) | | 2.31 (0.72, 6.54) |
| I am unable to bathe or dress myself | | 0.72 (0, 4) | 0.26 (0.08, 0.77) |
| Missing | 0.8 (0.23, 2) | 1.07 (0.5, 2.25) | 5.9 (3.16, 12.8) |
| **EQ-5D usual activities at 30 day follow-up** | | | |
| I have no problems in performing my usual activities | 0.75 (0.5, 0.91) | 1.04 (0.93, 1.18) | 1.14 (0.91, 1.53) |
| I have some problems in performing my usual activities | | 0.36 (0, 3.19) | 0.63 (0.28, 1.97) |
| I am unable to perform my usual activities | | 0.72 (0, 4) | 0.71 (0.17, 2.61) |
| Missing | 0.8 (0.23, 2) | 1.07 (0.5, 2.25) | 5.9 (3.16, 12.8) |
| **EQ-5D pain/discomfort at 30 day follow-up** | | | |
| I have no pain or discomfort | 1.19 (0.71, 3.32) | 1.12 (0.92, 1.37) | 1.07 (0.89, 1.42) |
| I have moderate pain or discomfort | 0.61 (0.16, 2.41) | 0.67 (0.23, 1.91) | 0.73 (0.34, 1.53) |
| I have extreme pain or discomfort | | | |
| Missing | 0.8 (0.23, 2) | 1.07 (0.5, 2.25) | 5.9 (3.16, 12.8) |
| **EQ-5D anxiety/depression at 30 day follow-up** | | | |
| I am not anxious or depressed | 0.85 (0.6, 0.95) | 1.13 (1.02, 1.35) | 0.98 (0.92, 1.09) |
| I am moderately anxious or depressed | | 0.28 (0, 1.71) | 1.22 (0.37, 3.33) |
| I am extremely anxious or depressed | | | |
| Missing | 0.8 (0.23, 2) | 1.07 (0.5, 2.25) | 5.9 (3.16, 12.8) |
| **Patient satisfaction** | | | |
| Very satisfied | 1.23 (0.84, 2.13) | 1.29 (1.09, 1.52) | 1.04 (0.83, 1.36) |
| Somewhat satisfied | 0.36 (0, 1.64) | 0.36 (0.07, 1.12) | 0.61 (0.29, 1.23) |
| Somewhat dissatisfied | | | 1.22 (0.38, 3.91) |
| Very dissatisfied | | | 2.23 (0.78, 4.78) |
| Missing | 2 (0.19, 7) | 0.5 (0, 2) | 11.5 (3.67, 28) |
| **Number of hospitalizations for this injury** | | | |
| Missing | 0.3 (0, 1.21) | 1.38 (0.5, 3.64) | 17 (5.33, 41) |
| **EQ-5D health state at discharge** | 1.08 (0.67, 1.75) | 1.12 (0.89, 1.12) | 1 (0.08, 15) |
| Missing | 2 (0.5, 6) | 1.19 (0.6, 2.45) | 4.8 (2.38, 11) |
| **EQ-5D health state at 30 day follow-up** | 0.83 (0.67, 1.33) | | 8.3 (0.74, 11.14) |
| Missing | 0.8 (0.23, 2) | 1.13 (0.53, 2.27) | 4.36 (2.43, 7.75) |
| **Cost of treatment** | 0.71 (0.14, 5) | 0.74 (0.2, 2.69) | 0.18 (0.04, 4) |
| Missing | 1.5 (0.66, 2.75) | 1.17 (0.6, 2.22) | 3.91 (2.37, 6.04) |

#### Table S7. Absolute and relative differences in outcomes after training, comparing standard care with ATLS(95% CI)

| Outcome | Absolute difference | Relative difference |
|---|---|---|
| **30 day mortality** | | |
| Yes | 12.2 (-8.05, 23.3) | 4.21 (1.36, 15.71) |
| No | -12 (-23, 6) | 0.88 (0.75, 1.1) |
| Missing | 24 (15.57, 39) | 13 (5.33, 40) |
| **24 hour mortality** | | |
| Yes | | |
| No | | |
| Missing | 5 (0, 13) | 3.5 (1.6, 14) |
| **In-hospital mortality** | | |
| Yes | 8.3 (-5.9, 18) | 3.24 (0.63, 13.64) |
| No | -8 (-18, 5) | 0.92 (0.82, 1.07) |
| Missing | 1 (-3.52, 5) | 2 (0.5, 5) |
| **Self-ambulatory at discharge** | | |
| Yes | -1 (-9, 2) | 0.99 (0.93, 1.03) |
| No | 1.5 (-2.2, 8.92) | |
| Missing | 20 (-23, 35) | 6 (0.1, 33) |
| **Return to work** | | |
| Yes | -5 (-30, 29) | 0.91 (0.6, 1.64) |
| No | 5 (-30, 30) | 1.12 (0.44, 2.41) |
| Missing | 47 (30, 63) | 6.22 (4.55, 53) |
| **Pulmonary complication** | | |
| Yes | | |
| No | | |
| Missing | 28 (5, 49) | 6.6 (4.33, 43) |
| **Septic complication** | | |
| Yes | 0.8 (-4.5, 8.26) | |
| No | -1 (-11, 2) | 0.99 (0.9, 1.03) |
| Missing | 28 (1, 50) | 6.6 (4.25, 43) |
| **Renal failure** | | |
| Yes | | |
| No | | |
| Missing | 27 (4, 48) | 6.4 (4.11, 42) |
| **Coagulopathy** | | |
| Yes | -4.3 (-16, 0) | |
| No | 4 (0, 16) | 1.04 (1, 1.19) |
| Missing | 27 (4, 48) | 6.4 (4.11, 42) |

| Outcome | Absolute difference | Relative difference |
|---|---|---|
| **Need for reexploration or resurgery** | | |
| Yes | 2.5 (0, 7.8) | |
| No | -3 (-8, -2) | 0.97 (0.92, 0.98) |
| Missing | 36 (-3.47, 56) | 7 (4.22, 48) |
| **Failure of conservative management** | | |
| No | 6 (2, 26) | 1.07 (1.02, 1.36) |
| Yes | -6.3 (-26, -2.4) | 0.21 (0, 2.28) |
| Missing | 41 (30, 56) | 14.67 (8.42, 55) |
| **EQ-5D mobility at discharge** | | |
| I have no problems in walking | -27 (-44, 31) | 0.7 (0.55, 1.52) |
| I have some problems in walking | 8 (-18.1, 23.91) | 1.73 (0.08, 12.79) |
| I am confined to bed | 19 (1.6, 30) | |
| Missing | 33 (-23, 50) | 4.3 (2.38, 20.5) |
| **EQ-5D self-care at discharge** | | |
| I have no problems with self-care | -17 (-46.52, 43) | 0.75 (0.48, 1.93) |
| I have some problems bathing or dressing myself | 20 (-24.9, 36.87) | 2.82 (0.2, 20.26) |
| I am unable to bathe or dress myself | -3 (-26.15, 19.7) | 0.86 (0.07, 7.29) |
| Missing | 33 (-23, 50) | 4.3 (2.38, 20.5) |
| **EQ-5D usual activities at discharge** | | |
| I have no problems in performing my usual activities | -14 (-49, 47) | 0.77 (0.45, 2.14) |
| I have some problems in performing my usual activities | 16 (-36.56, 39.71) | 1.73 (0.1, 12.04) |
| I am unable to perform my usual activities | -2 (-24.06, 14.32) | 0.88 (0, 5.17) |
| Missing | 33 (-23, 50) | 4.3 (2.38, 20.5) |
| **EQ-5D pain/discomfort at discharge** | | |
| I have no pain or discomfort | -25 (-52, 49) | 0.5 (0.25, 3.07) |
| I have moderate pain or discomfort | -15 (-49, -3) | 0.7 (0.26, 1.03) |
| I have extreme pain or discomfort | 40 (29.7, 55) | |
| Missing | 33 (-23, 50) | 4.3 (2.38, 20.5) |
| **EQ-5D anxiety/depression at discharge** | | |
| I am not anxious or depressed | 9 (2, 36) | 1.11 (1.03, 1.68) |
| I am moderately anxious or depressed | -10.2 (-39.9, -3.9) | 0.4 (0, 1.24) |
| I am extremely anxious or depressed | 1.7 (-1.5, 6.4) | |
| Missing | 33 (-23, 50) | 4.3 (2.38, 20.5) |
| **EQ-5D mobility at 30 day follow-up** | | |
| I have no problems in walking | -3 (-28, 30) | 0.96 (0.71, 1.43) |
| I have some problems in walking | -3 (-29, 22) | 0.85 (0, 5.21) |
| I am confined to bed | 5.9 (0.97, 15) | |
| Missing | 51 (34, 69) | 7.38 (5.4, 53) |
| **EQ-5D self-care at 30 day follow-up** | | |

| Outcome | Absolute difference | Relative difference |
|---|---|---|
| I have no problems with self-care | -1 (-25, 27) | 0.99 (0.74, 1.39) |
| I have some problems bathing or dressing myself | 5 (-20, 22) | 1.5 (0.83, 4.17) |
| I am unable to bathe or dress myself | -4.1 (-29, 1.5) | 0.59 (0, 2.6) |
| Missing | 51 (34, 69) | 7.38 (5.4, 53) |
| **EQ-5D usual activities at 30 day follow-up** | | |
| I have no problems in performing my usual activities | 6 (-21, 31) | 1.08 (0.79, 1.58) |
| I have some problems in performing my usual activities | -3 (-20.77, 16.16) | 0.8 (0, 11.66) |
| I am unable to perform my usual activities | -3.1 (-29, 5) | 0.69 (0, 3.76) |
| Missing | 51 (34, 69) | 7.38 (5.4, 53) |
| **EQ-5D pain/discomfort at 30 day follow-up** | | |
| I have no pain or discomfort | -1 (-18, 17) | 0.99 (0.8, 1.24) |
| I have moderate pain or discomfort | -1 (-20, 13) | 0.95 (0.25, 2.7) |
| I have extreme pain or discomfort | 2 (-1, 8.8) | |
| Missing | 51 (34, 69) | 7.38 (5.4, 53) |
| **EQ-5D anxiety/depression at 30 day follow-up** | | |
| I am not anxious or depressed | 10 (3, 36) | 1.12 (1.03, 1.58) |
| I am moderately anxious or depressed | -11.1 (-35.8, -2.9) | 0.26 (0, 1.11) |
| I am extremely anxious or depressed | 1 (-0.9, 5.1) | |
| Missing | 51 (34, 69) | 7.38 (5.4, 53) |
| **Patient satisfaction** | | |
| Very satisfied | -9 (-31, 30) | 0.89 (0.67, 1.46) |
| Somewhat satisfied | 1 (-15.8, 15.15) | 1.08 (0.09, 6.5) |
| Somewhat dissatisfied | 5.2 (-13, 15) | 2.24 (1, 4.12) |
| Very dissatisfied | 1.6 (-9.3, 9.53) | 1.38 (0.81, 3.95) |
| Missing | 19 (-21, 34.53) | 5.75 (0.11, 31.03) |
| **Number of hospitalizations for this injury** | | |
| Missing | 31 (21, 47) | 11.33 (6.8, 48) |
| **EQ-5D health state at discharge** | -15 (-84, 75.14) | 0.77 (0.06, 9.82) |
| Missing | 38 (24, 53) | 4.8 (3.23, 23) |
| **EQ-5D health state at 30 day follow-up** | 0 (-60, 50) | 1 (0.37, 2) |
| Missing | 53 (40, 73) | 7.62 (5.5, 53) |
| **Cost of treatment** | -2500 (-33500, 250) | 0.5 (0, 1.5) |
| Missing | 69 (-68, 98) | 4.29 (0.25, 9.3) |

#### Table S8. Absolute and relative differences in outcomes after training, comparing standard care with PTC(95% CI)

| Outcome | Absolute difference | Relative difference |
|---|---|---|
| **30 day mortality** | | |
| Yes | 11.1 (-5.3, 22.33) | 3.27 (0.61, 18.82) |
| No | -11 (-22.46, 4) | 0.88 (0.78, 1.05) |
| Missing | 14 (-24.07, 32) | 2.17 (0, 12) |
| **24 hour mortality** | | |
| Yes | | |
| No | | |
| Missing | -1 (-16, 4) | 0.88 (0, 3.33) |
| **In-hospital mortality** | | |
| Yes | 7.9 (-3.66, 16) | 2.93 (0.74, 17.86) |
| No | -8 (-24, 2) | 0.92 (0.8, 1.03) |
| Missing | 2 (-1, 6) | |
| **Self-ambulatory at discharge** | | |
| Yes | 2 (-1, 12) | 1.02 (1, 1.14) |
| No | -1.4 (-12, 0.7) | |
| Missing | 21 (-1, 45) | 8 (1.25, 46) |
| **Return to work** | | |
| Yes | -17 (-41, -4) | 0.76 (0.42, 0.98) |
| No | 17 (1, 41) | 1.57 (1.05, 6.48) |
| Missing | 40 (-36, 64) | 3.5 (0.18, 12.99) |
| **Pulmonary complication** | | |
| Yes | | |
| No | | |
| Missing | 23 (-22, 40) | 3.3 (0.12, 14.65) |
| **Septic complication** | | |
| Yes | -2.4 (-8.7, 0) | |
| No | 2 (0, 9) | 1.02 (1, 1.1) |
| Missing | 23 (-20, 41) | 3.3 (0.13, 15) |
| **Renal failure** | | |
| Yes | | |
| No | | |
| Missing | 22 (-20, 39.08) | 3.2 (0.13, 14.5) |
| **Coagulopathy** | | |
| Yes | 0 (-17.31, 4.2) | |
| No | 0 (-17, 0) | 1 (0.83, 1) |
| Missing | 22 (-20, 39.08) | 3.2 (0.13, 14.5) |

| Outcome | Absolute difference | Relative difference |
|---|---|---|
| **Need for reexploration or resurgery** | | |
| Yes | 0.9 (-5.32, 4.2) | |
| No | -1 (-5, 4) | 0.99 (0.95, 1.04) |
| Missing | 32 (-21.3, 51.96) | 4.2 (0.17, 17.75) |
| **Failure of conservative management** | | |
| No | 0 (-22, 8) | 1 (0.77, 1.09) |
| Yes | 0.1 (-8.4, 22) | 1.06 (0.26, 17) |
| Missing | 32 (-35.17, 54) | 3.67 (0, 20.5) |
| **EQ-5D mobility at discharge** | | |
| I have no problems in walking | -30 (-50, -1.99) | 0.67 (0.5, 1) |
| I have some problems in walking | 14 (0.84, 33) | 3.8 (2.06, 16.11) |
| I am confined to bed | 15.7 (-15.7, 28) | |
| Missing | 30 (-10, 51) | 3.31 (0.35, 13.38) |
| **EQ-5D self-care at discharge** | | |
| I have no problems with self-care | -35 (-52, -23) | 0.59 (0.44, 0.73) |
| I have some problems bathing or dressing myself | 22.7 (0, 41) | 3.73 (1.21, 20.62) |
| I am unable to bathe or dress myself | 12.3 (-5.5, 44.8) | 2.84 (0.88, 20) |
| Missing | 30 (-10, 51) | 3.31 (0.35, 13.38) |
| **EQ-5D usual activities at discharge** | | |
| I have no problems in performing my usual activities | -38 (-61, -23) | 0.55 (0.35, 0.69) |
| I have some problems in performing my usual activities | 29.7 (17, 46.5) | 4.58 (2.2, 26.47) |
| I am unable to perform my usual activities | 8.3 (-6, 33) | 2.24 (0.76, 18.12) |
| Missing | 30 (-10, 51) | 3.31 (0.35, 13.38) |
| **EQ-5D pain/discomfort at discharge** | | |
| I have no pain or discomfort | -40 (-62, -22) | 0.38 (0.21, 0.57) |
| I have moderate pain or discomfort | 5 (-16, 45) | 1.17 (0.69, 4.46) |
| I have extreme pain or discomfort | 35 (-36.75, 49) | |
| Missing | 30 (-10, 51) | 3.31 (0.35, 13.38) |
| **EQ-5D anxiety/depression at discharge** | | |
| I am not anxious or depressed | -3 (-40, 10) | 0.97 (0.59, 1.13) |
| I am moderately anxious or depressed | 1.8 (-11.4, 39.6) | 1.36 (0.43, 19.33) |
| I am extremely anxious or depressed | 1.7 (0, 6.4) | |
| Missing | 30 (-10, 51) | 3.31 (0.35, 13.38) |
| **EQ-5D mobility at 30 day follow-up** | | |
| I have no problems in walking | -21 (-50, -14) | 0.79 (0.5, 0.86) |
| I have some problems in walking | 17 (5, 50) | |
| I am confined to bed | 4.2 (-3.7, 12) | |
| Missing | 44 (-43.29, 69) | 3.93 (0.16, 15.75) |
| **EQ-5D self-care at 30 day follow-up** | | |

| Outcome | Absolute difference | Relative difference |
|---|---|---|
| I have no problems with self-care | -18 (-39, -10) | 0.81 (0.61, 0.9) |
| I have some problems bathing or dressing myself | 15 (7.4, 33) | |
| I am unable to bathe or dress myself | 2.5 (-7, 25.9) | 1.74 (0.58, 11.76) |
| Missing | 44 (-43.29, 69) | 3.93 (0.16, 15.75) |
| **EQ-5D usual activities at 30 day follow-up** | | |
| I have no problems in performing my usual activities | -14 (-46, 1) | 0.85 (0.52, 1.03) |
| I have some problems in performing my usual activities | 10.3 (-1, 41.2) | 7.06 (4.78, 23.89) |
| I am unable to perform my usual activities | 3.5 (-5.6, 24.9) | 2.03 (0.84, 11.76) |
| Missing | 44 (-43.29, 69) | 3.93 (0.16, 15.75) |
| **EQ-5D pain/discomfort at 30 day follow-up** | | |
| I have no pain or discomfort | -7 (-28, 6) | 0.92 (0.72, 1.09) |
| I have moderate pain or discomfort | 5 (-10, 26.65) | 1.36 (0.61, 14.5) |
| I have extreme pain or discomfort | 2 (0, 8.8) | |
| Missing | 44 (-43.29, 69) | 3.93 (0.16, 15.75) |
| **EQ-5D anxiety/depression at 30 day follow-up** | | |
| I am not anxious or depressed | -2 (-42, 7) | 0.98 (0.57, 1.09) |
| I am moderately anxious or depressed | 0.5 (-11, 42.2) | 1.15 (0.31, 27.27) |
| I am extremely anxious or depressed | 1 (0, 5.1) | |
| Missing | 44 (-43.29, 69) | 3.93 (0.16, 15.75) |
| **Patient satisfaction** | | |
| Very satisfied | -24 (-39, -17) | 0.74 (0.6, 0.84) |
| Somewhat satisfied | 8.3 (-1.09, 35.6) | 2.46 (1.09, 19.33) |
| Somewhat dissatisfied | 9.4 (4, 17) | |
| Very dissatisfied | 5.8 (0, 13) | |
| Missing | 20 (-1, 44) | 7.67 (1.25, 45) |
| **Number of hospitalizations for this injury** | | |
| Missing | 23 (-27, 42.97) | 3.09 (0, 15.5) |
| **EQ-5D health state at discharge** | -40 (-89, -25) | 0.56 (0.06, 0.83) |
| Missing | 29 (-32, 53) | 2.53 (0.18, 8.62) |
| **EQ-5D health state at 30 day follow-up** | -17 (-91, 5) | 0.83 (0.09, 1.1) |
| Missing | 44 (-47, 69.25) | 3.59 (0.14, 14.75) |
| **Cost of treatment** | 1000 (-5000, 27188.83) | 1.67 (0.4, 26.67) |
| Missing | 69 (-5, 98.46) | 4.29 (0.78, 10.67) |

#### Table S9. Absolute and relative differences in outcomes after training, comparing ATLS with PTC(95% CI)

| Outcome | Absolute difference | Relative difference |
|---|---|---|
| **30 day mortality** | | |
| Yes | -1.1 (-22, 11.22) | 0.78 (0, 4.44) |
| No | 1 (-12, 22) | 1.01 (0.88, 1.28) |
| Missing | -10 (-37, 9) | 0.17 (0, 3.17) |
| **24 hour mortality** | | |
| Yes | | |
| No | | |
| Missing | -6 (-15, -4) | 0.25 (0, 0.88) |
| **In-hospital mortality** | | |
| Yes | -0.4 (-17, 9.31) | 0.9 (0, 3.94) |
| No | 0 (-11, 15) | 1 (0.89, 1.19) |
| Missing | 1 (-2, 6) | |
| **Self-ambulatory at discharge** | | |
| Yes | 3 (0, 14) | 1.03 (1, 1.16) |
| No | -2.9 (-14, -1.4) | |
| Missing | 1 (-28, 25) | 1.33 (0, 13) |
| **Return to work** | | |
| Yes | -12 (-40, 7) | 0.83 (0.42, 1.16) |
| No | 12 (-9, 40) | 1.4 (0.86, 4.93) |
| Missing | -7 (-59, 9) | 0.56 (0.05, 2.11) |
| **Pulmonary complication** | | |
| Yes | | |
| No | | |
| Missing | -5 (-42, 5) | 0.5 (0, 2.2) |
| **Septic complication** | | |
| Yes | -3.2 (-8.2, -0.8) | |
| No | 3 (0, 8) | 1.03 (1, 1.09) |
| Missing | -5 (-41, 5) | 0.5 (0, 2.2) |
| **Renal failure** | | |
| Yes | | |
| No | | |
| Missing | -5 (-39, 5) | 0.5 (0, 2.2) |
| **Coagulopathy** | | |
| Yes | 4.3 (0, 16) | |
| No | -4 (-16, 0) | 0.96 (0.84, 1) |
| Missing | -5 (-39, 5) | 0.5 (0, 2.2) |

| Outcome | Absolute difference | Relative difference |
|---|---|---|
| **Need for reexploration or resurgery** | | |
| Yes | -1.6 (-7.7, 0) | |
| No | 2 (0, 8) | 1.02 (1, 1.09) |
| Missing | -4 (-46, 5) | 0.6 (0, 2.4) |
| **Failure of conservative management** | | |
| No | -6 (-26, -1) | 0.94 (0.73, 0.99) |
| Yes | 6.4 (0.9, 26.4) | 5 (2.6, 27.14) |
| Missing | -9 (-52, 9) | 0.25 (0, 3) |
| **EQ-5D mobility at discharge** | | |
| I have no problems in walking | -3 (-37.88, 35.16) | 0.97 (0.61, 1.59) |
| I have some problems in walking | 6 (-14, 25.04) | 2.2 (0.45, 14.67) |
| I am confined to bed | -3.3 (-26, 3.6) | |
| Missing | -3 (-51, 9) | 0.77 (0.11, 2.25) |
| **EQ-5D self-care at discharge** | | |
| I have no problems with self-care | -18 (-57, 21) | 0.79 (0.41, 1.49) |
| I have some problems bathing or dressing myself | 2.7 (-30, 29.35) | 1.33 (0, 8) |
| I am unable to bathe or dress myself | 15.3 (4, 42.8) | 3.28 (1.16, 20) |
| Missing | -3 (-51, 9) | 0.77 (0.11, 2.25) |
| **EQ-5D usual activities at discharge** | | |
| I have no problems in performing my usual activities | -24 (-63, 10.64) | 0.72 (0.28, 1.39) |
| I have some problems in performing my usual activities | 13.7 (-21.15, 56) | 2.65 (0.74, 23.16) |
| I am unable to perform my usual activities | 10.3 (-1, 50.3) | 2.54 (1.09, 18.33) |
| Missing | -3 (-51, 9) | 0.77 (0.11, 2.25) |
| **EQ-5D pain/discomfort at discharge** | | |
| I have no pain or discomfort | -15 (-53, 33.45) | 0.77 (0.3, 2.35) |
| I have moderate pain or discomfort | 20 (3, 49) | 1.67 (1.12, 4.46) |
| I have extreme pain or discomfort | -5 (-49, 5.8) | |
| Missing | -3 (-51, 9) | 0.77 (0.11, 2.25) |
| **EQ-5D anxiety/depression at discharge** | | |
| I am not anxious or depressed | -12 (-43, -4) | 0.87 (0.55, 0.96) |
| I am moderately anxious or depressed | 12 (1.7, 42.8) | 3.4 (1.17, 22.31) |
| I am extremely anxious or depressed | 0 (-6.8, 0) | |
| Missing | -3 (-51, 9) | 0.77 (0.11, 2.25) |
| **EQ-5D mobility at 30 day follow-up** | | |
| I have no problems in walking | -18 (-55, -4) | 0.82 (0.45, 0.98) |
| I have some problems in walking | 20 (11.9, 55) | |
| I am confined to bed | -1.7 (-12, 1.7) | |
| Missing | -7 (-65, 8) | 0.53 (0.05, 2.14) |
| **EQ-5D self-care at 30 day follow-up** | | |

| Outcome | Absolute difference | Relative difference |
|---|---|---|
| I have no problems with self-care | -17 (-38, -8) | 0.82 (0.61, 0.94) |
| I have some problems bathing or dressing myself | 10 (-6, 30) | |
| I am unable to bathe or dress myself | 6.6 (2.4, 24.3) | 2.94 (1.65, 13.89) |
| Missing | -7 (-65, 8) | 0.53 (0.05, 2.14) |
| **EQ-5D usual activities at 30 day follow-up** | | |
| I have no problems in performing my usual activities | -20 (-46, -12) | 0.79 (0.53, 0.88) |
| I have some problems in performing my usual activities | 13.3 (5, 38) | 8.82 (9.31, 21.11) |
| I am unable to perform my usual activities | 6.6 (1.8, 24) | 2.94 (1.7, 13.89) |
| Missing | -7 (-65, 8) | 0.53 (0.05, 2.14) |
| **EQ-5D pain/discomfort at 30 day follow-up** | | |
| I have no pain or discomfort | -6 (-29, 7) | 0.93 (0.69, 1.11) |
| I have moderate pain or discomfort | 6 (-8, 28) | 1.43 (0.67, 12) |
| I have extreme pain or discomfort | 0 (-6.6, 0) | |
| Missing | -7 (-65, 8) | 0.53 (0.05, 2.14) |
| **EQ-5D anxiety/depression at 30 day follow-up** | | |
| I am not anxious or depressed | -12 (-34, -3) | 0.88 (0.65, 0.98) |
| I am moderately anxious or depressed | 11.6 (-0.06, 34.4) | 4.41 (0.87, 23.75) |
| I am extremely anxious or depressed | 0 (-4.8, 0) | |
| Missing | -7 (-65, 8) | 0.53 (0.05, 2.14) |
| **Patient satisfaction** | | |
| Very satisfied | -15 (-35, 8) | 0.84 (0.65, 1.16) |
| Somewhat satisfied | 7.3 (-2.4, 26.7) | 2.28 (0.93, 12.14) |
| Somewhat dissatisfied | 4.2 (-7.79, 15.31) | |
| Very dissatisfied | 4.2 (-3, 17) | |
| Missing | 1 (-27.21, 24) | 1.33 (0, 13) |
| **Number of hospitalizations for this injury** | | |
| Missing | -8 (-44, 8) | 0.27 (0, 2.75) |
| **EQ-5D health state at discharge** | -25 (-85, 15) | 0.72 (0.06, 1.39) |
| Missing | -9 (-55, 10) | 0.53 (0.08, 1.92) |
| **EQ-5D health state at 30 day follow-up** | -17 (-91, 0) | 0.83 (0.09, 1) |
| Missing | -9 (-67, 9) | 0.47 (0.04, 2.25) |
| **Cost of treatment** | 3500 (0, 33000) | 3.33 (1, 55.56) |
| Missing | 0 (-95.49, 70) | 1 (0.11, 4.45) |

#### Table S10. Absolute and relative differences in changes from baseline for all outcomes, comparing standard care with ATLS(95% CI)

| Outcome | Absolute difference | Relative difference |
|---|---|---|
| **30 day mortality** | | |
| Yes | 9.6 (-25.64, 31.44) | |
| No | -9 (-31, 26) | |
| Missing | 23 (-9.51, 43) | |
| **24 hour mortality** | | |
| Yes | -2.4 (-9.8, 0) | |
| No | 2 (-3, 10) | |
| Missing | 7 (-1, 18) | |
| **In-hospital mortality** | | |
| Yes | 3.4 (-16.6, 21.53) | |
| No | -3 (-22, 16) | |
| Missing | 1 (-3.52, 5) | 2 (0.5, 5) |
| **Self-ambulatory at discharge** | | |
| Yes | -4 (-38, 4) | |
| No | 4.7 (-3.7, 38) | |
| Missing | 22 (-31, 39.93) | |
| **Return to work** | | |
| Yes | 37 (18, 86) | |
| No | -37 (-86, -19) | |
| Missing | 45 (-39, 71) | |
| **Pulmonary complication** | | |
| Yes | 0 (-12, 0) | |
| No | 0 (-10, 2) | |
| Missing | 32 (-33, 51.17) | |
| **Septic complication** | | |
| Yes | -1.7 (-12.85, 5.17) | |
| No | 1 (-8, 10) | |
| Missing | 33 (-36, 53.18) | |
| **Renal failure** | | |
| Yes | | |
| No | | |
| Missing | 31 (-34, 50.16) | |
| **Coagulopathy** | | |
| Yes | -4.3 (-16, 0) | |

| Outcome | Absolute difference | Relative difference |
|---|---|---|
| No | 4 (0, 16) | |
| Missing | 32 (-34, 51.29) | |
| **Need for reexploration or resurgery** | | |
| Yes | 2.5 (-3.8, 10.29) | |
| No | -3 (-11, 1) | |
| Missing | 39 (-43, 63.27) | 20.5 (7.8, 58) |
| **Failure of conservative management** | | |
| No | 6 (0, 26) | |
| Yes | -6.3 (-26, -1.5) | |
| Missing | 49 (33, 66) | |
| **EQ-5D mobility at discharge** | | |
| I have no problems in walking | -22 (-78, -6) | |
| I have some problems in walking | -1.7 (-26.99, 20.5) | |
| I am confined to bed | 23 (7.18, 63) | |
| Missing | 35 (-51.8, 60.45) | |
| **EQ-5D self-care at discharge** | | |
| I have no problems with self-care | -18 (-56, 6) | |
| I have some problems bathing or dressing myself | 22 (-5, 59) | |
| I am unable to bathe or dress myself | -4 (-42.7, 17.31) | |
| Missing | 35 (-51.8, 60.45) | |
| **EQ-5D usual activities at discharge** | | |
| I have no problems in performing my usual activities | -18 (-64, 15) | |
| I have some problems in performing my usual activities | 13 (-21.42, 46.56) | |
| I am unable to perform my usual activities | 5 (-12.9, 45) | |
| Missing | 35 (-51.8, 60.45) | |
| **EQ-5D pain/discomfort at discharge** | | |
| I have no pain or discomfort | -25 (-66.11, 51) | |
| I have moderate pain or discomfort | 11 (-43, 52.7) | |
| I have extreme pain or discomfort | 14 (-8, 37.6) | |
| Missing | 35 (-51.8, 60.45) | |
| **EQ-5D anxiety/depression at discharge** | | |
| I am not anxious or depressed | -20 (-73, -4) | 0.39 (-30, 1.56) |
| I am moderately anxious or depressed | 21.8 (6, 68) | 0.34 (-264, 1.29) |
| I am extremely anxious or depressed | -0.9 (-9.42, 5.6) | |
| Missing | 35 (-51.8, 60.45) | |
| **EQ-5D mobility at 30 day follow-up** | | |
| I have no problems in walking | -1 (-42, 28) | |

| Outcome | Absolute difference | Relative difference |
|---|---|---|
| I have some problems in walking | 1 (-33, 27) | |
| I am confined to bed | -0.1 (-29.57, 36.4) | |
| Missing | 51 (-40, 74) | |
| **EQ-5D self-care at 30 day follow-up** | | |
| I have no problems with self-care | 11 (-6, 59) | |
| I have some problems bathing or dressing myself | 15.5 (-8.77, 58.1) | |
| I am unable to bathe or dress myself | -27.1 (-60.9, -19.65) | |
| Missing | 51 (-40, 74) | |
| **EQ-5D usual activities at 30 day follow-up** | | |
| I have no problems in performing my usual activities | 35 (14, 76) | |
| I have some problems in performing my usual activities | -22 (-64, -9.9) | |
| I am unable to perform my usual activities | -12.8 (-38.4, -6.2) | |
| Missing | 51 (-40, 74) | |
| **EQ-5D pain/discomfort at 30 day follow-up** | | |
| I have no pain or discomfort | -8 (-74.56, 15) | |
| I have moderate pain or discomfort | 6 (-18, 58.86) | |
| I have extreme pain or discomfort | 2 (-6.57, 11.06) | |
| Missing | 51 (-40, 74) | |
| **EQ-5D anxiety/depression at 30 day follow-up** | | |
| I am not anxious or depressed | 13 (-14, 45) | |
| I am moderately anxious or depressed | -14.3 (-39.4, 1.3) | |
| I am extremely anxious or depressed | 1 (-5.8, 9.6) | |
| Missing | 51 (-40, 74) | |
| **Patient satisfaction** | | |
| Very satisfied | -12 (-45.57, 27) | |
| Somewhat satisfied | 14 (3.16, 53.3) | 0.39 (-27.05, 2.55) |
| Somewhat dissatisfied | -2.5 (-17.13, 10.9) | |
| Very dissatisfied | -1 (-21.48, 17.3) | |
| Missing | 19 (-29, 36) | |
| **Number of hospitalizations for this injury** | 0 (-1, -1) | |
| Missing | 39 (29, 57) | |
| **EQ-5D health state at discharge** | -5 (-68.92, 60) | |
| Missing | 33 (-38, 62.24) | |
| **EQ-5D health state at 30 day follow-up** | 90 (86, 116) | |
| Missing | 49 (-35, 77) | |
| **Cost of treatment** | -9500 (-74630.2, 10549.64) | |
| Missing | 60 (-66, 95) | 9.57 (0.05, 94) |

#### Table S11. Absolute and relative differences in changes from baseline for all outcomes, comparing standard care with PTC(95% CI)

| Outcome | Absolute difference | Relative difference |
|---|---|---|
| **30 day mortality** | | |
| Yes | 18.5 (14.1, 44.3) | |
| No | -18 (-44, -14) | |
| Missing | 19 (-14.83, 37) | |
| **24 hour mortality** | | |
| Yes | -2.4 (-9.8, 0) | |
| No | 2 (0, 10) | |
| Missing | 4 (-10, 12) | |
| **In-hospital mortality** | | |
| Yes | 10 (3.8, 25.8) | |
| No | -10 (-26, -5) | |
| Missing | 2 (-1, 6) | |
| **Self-ambulatory at discharge** | | |
| Yes | 3 (-10, 40.7) | |
| No | -2.4 (-40.06, 10.2) | |
| Missing | 27 (15.73, 49) | |
| **Return to work** | | |
| Yes | 16 (-61, 54) | |
| No | -16 (-56, 57.88) | |
| Missing | 43 (-6, 69.51) | |
| **Pulmonary complication** | | |
| Yes | 2.2 (0, 10) | |
| No | -2 (-10, 0) | |
| Missing | 32 (-3, 52) | |
| **Septic complication** | | |
| Yes | -0.5 (-7.44, 7.44) | |
| No | 0 (-9, 5) | |
| Missing | 34 (-2, 55) | |
| **Renal failure** | | |
| Yes | | |
| No | | |
| Missing | 32 (-2, 51) | |
| **Coagulopathy** | | |
| Yes | 0 (-17.31, 4.2) | |

| Outcome | Absolute difference | Relative difference |
|---|---|---|
| No | 0 (-17, 0) | |
| Missing | 33 (-2, 52) | |
| **Need for reexploration or resurgery** | | |
| Yes | 3 (-0.1, 10.7) | |
| No | -3 (-11, -1) | |
| Missing | 41 (-1, 64) | |
| **Failure of conservative management** | | |
| No | -2 (-27, 4) | |
| Yes | 2.3 (-4.1, 27.3) | |
| Missing | 41 (-42, 64) | |
| **EQ-5D mobility at discharge** | | |
| I have no problems in walking | -9 (-45, 37.2) | |
| I have some problems in walking | 8 (-10.76, 43.6) | |
| I am confined to bed | -0.4 (-71.7, 22) | |
| Missing | 44 (33, 74) | |
| **EQ-5D self-care at discharge** | | |
| I have no problems with self-care | -14 (-67.77, 24) | |
| I have some problems bathing or dressing myself | 19.7 (-9.7, 62.13) | |
| I am unable to bathe or dress myself | -6.4 (-47, 21) | |
| Missing | 44 (33, 74) | |
| **EQ-5D usual activities at discharge** | | |
| I have no problems in performing my usual activities | -16 (-72.97, 22) | |
| I have some problems in performing my usual activities | 16.7 (-15, 65.46) | |
| I am unable to perform my usual activities | 0.1 (-34, 32.74) | |
| Missing | 44 (33, 74) | |
| **EQ-5D pain/discomfort at discharge** | | |
| I have no pain or discomfort | -39 (-78.7, -13) | |
| I have moderate pain or discomfort | 27 (0, 61) | |
| I have extreme pain or discomfort | 11.4 (-3.77, 36.29) | |
| Missing | 44 (33, 74) | |
| **EQ-5D anxiety/depression at discharge** | | |
| I am not anxious or depressed | -9 (-45.55, 37.45) | 0.59 (0, 9.18) |
| I am moderately anxious or depressed | 7.8 (-42.26, 43.34) | 0.59 (0.03, 12.41) |
| I am extremely anxious or depressed | 1.5 (-2, 13.5) | |
| Missing | 44 (33, 74) | |
| **EQ-5D mobility at 30 day follow-up** | | |
| I have no problems in walking | 5 (-35, 49) | |

| Outcome | Absolute difference | Relative difference |
|---|---|---|
| I have some problems in walking | 11 (-16, 83) | |
| I am confined to bed | -16.5 (-82.9, 0) | |
| Missing | 48 (-7, 76.59) | |
| **EQ-5D self-care at 30 day follow-up** | | |
| I have no problems with self-care | 2 (-65.02, 32) | |
| I have some problems bathing or dressing myself | 13.2 (-14.34, 87.99) | |
| I am unable to bathe or dress myself | -15.8 (-44.79, 21.8) | |
| Missing | 48 (-7, 76.59) | |
| **EQ-5D usual activities at 30 day follow-up** | | |
| I have no problems in performing my usual activities | 6 (-70, 32) | |
| I have some problems in performing my usual activities | -4 (-21.6, 47.5) | |
| I am unable to perform my usual activities | -1.5 (-17, 36.26) | |
| Missing | 48 (-7, 76.59) | |
| **EQ-5D pain/discomfort at 30 day follow-up** | | |
| I have no pain or discomfort | -4 (-61.12, 34.64) | |
| I have moderate pain or discomfort | 0 (-48, 45) | |
| I have extreme pain or discomfort | 4.3 (2.1, 12.9) | |
| Missing | 48 (-7, 76.59) | |
| **EQ-5D anxiety/depression at 30 day follow-up** | | |
| I am not anxious or depressed | -13 (-60, 8) | |
| I am moderately anxious or depressed | 9.3 (-11.4, 60.2) | |
| I am extremely anxious or depressed | 3.3 (2, 12.9) | |
| Missing | 48 (-7, 76.59) | |
| **Patient satisfaction** | | |
| Very satisfied | -18 (-66, 6) | |
| Somewhat satisfied | 1.3 (-41.15, 30.03) | |
| Somewhat dissatisfied | 5.6 (-3.77, 19.4) | |
| Very dissatisfied | 11 (4.3, 25) | |
| Missing | 24 (15, 48) | |
| **Number of hospitalizations for this injury** | 0 (-1, -1) | |
| Missing | 29 (-33.53, 52) | |
| **EQ-5D health state at discharge** | -10 (-90, 39) | |
| Missing | 35 (-3, 59) | |
| **EQ-5D health state at 30 day follow-up** | 73 (15, 114) | |
| Missing | 45 (-8, 73.22) | |
| **Cost of treatment** | -10975 (-81672.96, 7582.31) | |

| Outcome | Absolute difference | Relative difference |
| --- | --- | --- |
| Missing | 64 (-2, 103) | |

#### Table S12. Absolute and relative differences in changes from baseline for all outcomes, comparing ATLS with PTC(95% CI)

| Outcome | Absolute difference | Relative difference |
|---|---|---|
| **30 day mortality** | | |
| Yes | 8.9 (-12.57, 32.12) | |
| No | -9 (-32.63, 12) | |
| Missing | -4 (-37, 7) | |
| **24 hour mortality** | | |
| Yes | 0 (-12, 0) | |
| No | 0 (-11, 0) | |
| Missing | -3 (-15, 3) | |
| **In-hospital mortality** | | |
| Yes | 6.6 (-4.2, 24.9) | |
| No | -7 (-25, 2) | |
| Missing | 1 (-2, 6) | |
| **Self-ambulatory at discharge** | | |
| Yes | 7 (0, 41) | |
| No | -7.1 (-41.1, -0.7) | |
| Missing | 5 (-25, 33) | |
| **Return to work** | | |
| Yes | -21 (-89, 10) | |
| No | 21 (-12, 89) | |
| Missing | -2 (-66, 41) | |
| **Pulmonary complication** | | |
| Yes | 2.2 (0, 10) | |
| No | -2 (-10, 0) | |
| Missing | 0 (-47, 36) | |
| **Septic complication** | | |
| Yes | 1.2 (-3.75, 13.86) | |
| No | -1 (-14, 3) | |
| Missing | 1 (-44.28, 39) | |
| **Renal failure** | | |
| Yes | | |
| No | | |
| Missing | 1 (-41.9, 37) | |
| **Coagulopathy** | | |
| Yes | 4.3 (0, 16) | |

| Outcome | Absolute difference | Relative difference |
|---|---|---|
| No | -4 (-16, 0) | |
| Missing | 1 (-42.61, 38) | |
| **Need for reexploration or resurgery** | | |
| Yes | 0.5 (-6.1, 5.2) | |
| No | 0 (-5, 6) | |
| Missing | 2 (-46, 49) | |
| **Failure of conservative management** | | |
| No | -8 (-28, -2) | |
| Yes | 8.6 (2.1, 28.5) | |
| Missing | -8 (-67, 9) | |
| **EQ-5D mobility at discharge** | | |
| I have no problems in walking | 13 (-12, 72) | |
| I have some problems in walking | 9.7 (-7.7, 45) | |
| I am confined to bed | -23.4 (-57.5, -1.76) | |
| Missing | 9 (-39, 51) | |
| **EQ-5D self-care at discharge** | | |
| I have no problems with self-care | 4 (-32.19, 58.2) | |
| I have some problems bathing or dressing myself | -2.3 (-50.44, 25.2) | |
| I am unable to bathe or dress myself | -2.4 (-38, 33) | |
| Missing | 9 (-39, 51) | |
| **EQ-5D usual activities at discharge** | | |
| I have no problems in performing my usual activities | 2 (-36, 56.83) | |
| I have some problems in performing my usual activities | 3.7 (-34.93, 48) | |
| I am unable to perform my usual activities | -4.9 (-51.52, 21) | |
| Missing | 9 (-39, 51) | |
| **EQ-5D pain/discomfort at discharge** | | |
| I have no pain or discomfort | -14 (-65.85, 40) | |
| I have moderate pain or discomfort | 16 (-23, 70) | |
| I have extreme pain or discomfort | -2.6 (-30, 6.85) | |
| Missing | 9 (-39, 51) | |
| **EQ-5D anxiety/depression at discharge** | | |
| I am not anxious or depressed | 11 (-15, 70) | 1.5 (0.5, 35) |
| I am moderately anxious or depressed | -14 (-67.3, 8.2) | 1.74 (0.56, 28.5) |
| I am extremely anxious or depressed | 2.4 (0, 11.4) | |
| Missing | 9 (-39, 51) | |
| **EQ-5D mobility at 30 day follow-up** | | |
| I have no problems in walking | 6 (-34, 57) | 1.86 (0.3, 44) |

| Outcome | Absolute difference | Relative difference |
|---|---|---|
| I have some problems in walking | 10 (-16, 87.6) | |
| I am confined to bed | -16.4 (-101.8, 2.94) | |
| Missing | -3 (-72, 40) | |
| **EQ-5D self-care at 30 day follow-up** | | |
| I have no problems with self-care | -9 (-93, 16) | |
| I have some problems bathing or dressing myself | -2.3 (-39.42, 30.96) | |
| I am unable to bathe or dress myself | 11.3 (-7.7, 62.3) | |
| Missing | -3 (-72, 40) | |
| **EQ-5D usual activities at 30 day follow-up** | | |
| I have no problems in performing my usual activities | -29 (-66, 7) | |
| I have some problems in performing my usual activities | 18 (-1.3, 44.9) | |
| I am unable to perform my usual activities | 11.3 (1.1, 34.5) | |
| Missing | -3 (-72, 40) | |
| **EQ-5D pain/discomfort at 30 day follow-up** | | |
| I have no pain or discomfort | 4 (-39.62, 56.63) | |
| I have moderate pain or discomfort | -6 (-62.71, 31) | |
| I have extreme pain or discomfort | 2.3 (0, 12) | |
| Missing | -3 (-72, 40) | |
| **EQ-5D anxiety/depression at 30 day follow-up** | | |
| I am not anxious or depressed | -26 (-49, -17.47) | |
| I am moderately anxious or depressed | 23.6 (12.2, 46) | |
| I am extremely anxious or depressed | 2.3 (0, 11) | |
| Missing | -3 (-72, 40) | |
| **Patient satisfaction** | | |
| Very satisfied | -6 (-47.26, 25) | |
| Somewhat satisfied | -12.7 (-57.2, 2.75) | |
| Somewhat dissatisfied | 8.1 (0.9, 30) | |
| Very dissatisfied | 12 (4.2, 26) | |
| Missing | 5 (-22, 30) | |
| **Number of hospitalizations for this injury** | 0 (-1, -0.5) | |
| Missing | -10 (-55, 11) | |
| **EQ-5D health state at discharge** | -5 (-78.91, 41.66) | |
| Missing | 2 (-44.02, 42) | |
| **EQ-5D health state at 30 day follow-up** | -17 (-109, 20) | |
| Missing | -4 (-71, 18) | |
| **Cost of treatment** | -1475 (-33639.83, 29927.2) | |

| Outcome | Absolute difference | Relative difference |
| --- | --- | --- |
| Missing | 4 (-75, 76) | |

### S13 Protocol Deviations

#### Trial Registration

We intended to register our trial with Clinical Trials Registry - India and will do so with the full-scale trial.

#### Outcomes across subgroups

Because of small numbers in the pre-specified subgroups men, women, blunt multisystem trauma, penetrating trauma, shock, severe traumatic brain injury and elderly we decided to report only descriptive data on these subgroups.

#### Number of Participating Centres

We recruited seven centres instead of six and therefore assigned two centres each to the intervention arms and three centres to the control arm.

#### Resident Participants

We included emergency medicine residents in addition to surgical residents.

#### Periodic surveys of residents

We did not distribute periodic surveys to the participating residents but discussed challenges and suggestions that they had regarding the scheduling or implementation of the training programs.

#### Follow up of residents

We stated that resident participants would be followed up 30 days after training, but revised this to follow them up after the end of the study period.

#### Data collection from records

We decided to extract data from medical records only for a convenience sample of patients to reduce the research officers' workload.

#### Selection of units for training

We planned to use simple random sampling to select units if there were more than two eligible units in a hospital but instead the hospital principal investigator decided which units to train.

#### Timing of resident consent

We had initially planned to ask residents for consent before randomisation, but because of logistical issues the units were only finalised after the hospitals had been randomised. Residents were therefore approached for consent after randomisation but before training.

#### Analysis level of feasibility outcomes

We had planned to analyse feasibility outcomes on both an overall and individual cluster level, but we only analysed them on an overall level, because the sample sizes in individual clusters were too small to generate meaningful results.
